# Lifestyle modifies the diabetes-related metabolic risk, conditional on individual genetic differences

**DOI:** 10.1101/2020.11.22.20236505

**Authors:** Jisu Shin, Xuan Zhou, Joanne Tan, Elina Hyppönen, Beben Benyamin, S Hong Lee

## Abstract

**Background:** Metabolic syndrome is a group of heritable metabolic traits that are highly associated with type 2 diabetes (T2DM). Classical interventions to T2DM include individual self-management of environmental risk factors such as improving diet quality, increasing physical activity and reducing smoking and alcohol consumptions, which decreases the risk of developing metabolic syndrome. However, it is poorly understood how the phenotypes of diabetes-related metabolic traits change with respect to lifestyle modifications at the individual level.

**Methods:** In this study, we applied a whole-genome genotype-by-environment (GxE) interaction approach to describe how intermediate traits reflecting metabolic risk are affected by genetic variations and how this genetic risk can interact with lifestyle, which can vary, conditional on individual genetic differences. In the analysis, we used 12 diabetes-related metabolic traits and eight lifestyle covariates from the UK Biobank comprising 288,837 white British participants genotyped for 1,133,273 genome-wide single nucleotide polymorphisms.

**Findings:** We found 17 GxE interactions, of which four modulated BMI and the others distributed across other traits. Modulation of genetic effects by physical activity was seen for four traits (glucose, HbA1c, C-reactive protein, systolic blood pressure), and by alcohol and smoking for three (BMI, glucose, waist-hip ratio; and BMI, diastolic and systolic blood pressure, respectively). We also found a number of significant phenotypic modulations by the lifestyle covariates, which were not attributed to the genetic effects in the model. Overall, modulation in the metabolic risk in response to the level of lifestyle covariates was clearly observed, and its direction and magnitude were varied depending on individual differences. We also showed that the metabolic risk inferred by our model was notably higher in T2DM prospective cases than controls.

**Interpretation:** Our findings highlight the importance of individual genetic differences in the prevention and management of diabetes and suggest that the one-size-fits-all approach may not benefit all.

**Funding:** This study has been supported by the Australian Research Council (DP 190100766, FT 160100229).

## Introduction

Diabetes mellitus is a metabolic disease normally caused by high blood glucose levels, which can lead to complications in kidneys, eyes and the nervous system ^1^. Currently, it is one of the top ten leading causes of death in the world ^2^, highlighting the importance of improved strategies on prevention and management. Type 2 diabetes mellitus (T2DM), which accounts for over 90% of all cases of diabetes ^3^, is known to be more polygenic than other types of diabetes ^4,5^. T2DM is often comorbid with other complex diseases such as cardiovascular diseases ^6,7^, and metabolic syndrome is highly associated with increasing the risk for both T2DM and cardiovascular diseases ^8,9^. Metabolic syndrome is a group of traits that causes metabolic diseases such as diabetes. Diabetes-related metabolic traits can include glucose, Haemoglobin A1c (HbA1c), C-reactive protein (CRP), body mass index (BMI), cholesterols and blood pressure (BP) ^10^.

Metabolic abnormalities and diabetes risk are affected by genetic factors ^11^. A recent genome-wide association study (GWAS) has identified 143 genetic variants associated with T2DM that have shed light on the aetiology of the disease 12. However, the identified genetic variants explain only a small proportion of phenotypic variance ^12-14^, which is unlikely to accurately predict the individual genetic (or polygenic) risk of T2DM in the early life stage ^15^. A whole-genome approach using all common single nucleotide polymorphisms (SNPs), instead of using a few genome-wide significant SNPs, has been proposed as a new promising approach for polygenic risk prediction ^16-18^. Recently, it has been shown that the accuracy of polygenic risk prediction can be increased further when using advanced statistical models and designs ^19-22^.

In addition to the genetic factors, T2DM risks are also increased by environmental conditions such as unhealthy diet and physical inactivity ^23^. Therefore, T2DM preventions and interventions include improving diet quality, increasing physical activity, and reducing smoking and alcohol consumptions. These interventions are often uniformly recommended for people with high metabolic risk irrespective of their response to these interventions. However, this one-size-fits-all approach may be inefficient because it does not consider individual genetic differences ^24-26^. In fact, it is little known how the T2DM risk in response to lifestyle modification can vary with respect to individuals’ genotypes, i.e. genotype-by-environment (GxE) interaction, and it is poorly understood how the information of GxE interaction can be incorporated in a T2DM intervention. It is unlikely that the changed genetic effects by lifestyle modification are in the same direction and magnitude for all people, therefore, the lifestyle modification should be tailored to each individual, considering individual genetic difference, i.e. personalised intervention.

In this study, we applied a whole-genome approach ^27^ to estimate the genetic and non-genetic effects on 12 diabetes-related metabolic traits modulated by eight lifestyle covariates. We show that the direction and magnitude of metabolic risk in response to the level of lifestyle covariates vary depending on individual genetic differences, i.e. phenotypic plasticity of the risk. We also showed that the predicted metabolic risk of T2DM prospective cases is significantly higher than controls. We conclude that a paradigm shift in intervention approaches for T2DM is required to account for individual differences, which will be realised as a precision medicine afforded by the increased availability of genomic data (e.g., UK biobank) and advanced computational models. This novel approach will enable more accurate treatments and preventions of T2DM.

## Methods

### Ethical Statement

UK Biobank’s scientific protocol and operational procedures were reviewed and approved by the North West Multi-Centre Research Ethics Committee (MREC), National Information Governance Board for Health & Social Care (NIGB), and Community Health Index Advisory Group (CHIAG). The access of the UK Biobank data was approved by the UK Biobank based on the application 14575 (“Whole-genome approaches for dissecting (shared) genetic architecture and individual risk prediction of complex traits in human populations”). Research Ethics was approved by the University of South Australia Human Research Ethics Committee (HREC).

### Participants

The UK Biobank consist of more than 500,000 individuals, recruited 22 assessment centres across the UK between March 2006 and Oct 2010 ^28^. The participants were recruited when they were between 37 to 73 years old ^29^, and all information used in this study are derived from information collected during the baseline survey.

### Phenotypic Data

As outcomes we used the information of diabetes-related metabolic traits including glucose, HbA1c, total cholesterol (TC), LDL cholesterol (LDL), HDL cholesterol (HDL), CRP, sodium and potassium in urine, BMI, waist-hip-ratio (WHR), and systolic BP and diastolic BP, with details on assay and measures given in supplementary material (Note S1). Lifestyle covariates were obtained by self-report and included age at recruitment, alcohol intake frequency (ALC), smoking status (SMK), metabolic equivalent task (MET) minutes per week for walking, moderate, vigorous, total activities and healthy dietary scores (Note S2).

### Genotypic Data

This study used the UK Biobank genotypic data, which comprise of 92,693,895 SNPs genotyped from 488,377 participants. In preliminary quality controls, we excluded individuals and SNPs that did not meet the following criteria from the UK Biobank data. At the individual level, we excluded individuals who were not white British (to reduce the effect of population stratification), have a missing rate ≥ 5%, have a gender mis-matched between self-reported and genetic data, and with putative sex chromosome aneuploidy. One individual from a pair, which has a genomic relationship larger than 0.05, was randomly selected and excluded. Furthermore, individuals who were population outliers (i.e. not within ± 6 standard deviations from the first and second principal components) were excluded. At the SNPs level, SNPs with an information score less than 0.6, with SNP missing rate less than 95%, with Hardy-Weinberg equilibrium P-value less than 0.0001 and with minor allele frequency less than 1% were excluded. Duplicated SNPs were also removed. The ratio discordant SNPs between the initial and second released individuals in the UK Biobank data was calculated, and additional 29 individuals who had a discordance rate more than 0.05 were excluded. We only used HapMap3 SNPs from the quality-controlled data, which are of high quality and well calibrated to dissect the genetic architecture of complex diseases ^30,31^. After quality controls, the cleaned data include 1,133,273 SNPs and 288,837 participants.

Due to the usage of the individual level of genotypic data, which demands high computational resources, we further divided samples into six groups. Of the total samples, 91,472 individuals from the first release of these selected individuals, divided into two groups. Meanwhile, 197,365 individuals from the second release were divided into four groups. Meta-analysed estimates and p-values across the six groups were reported.

### Data Analysis

#### Adjustment for the phenotypes of main traits and lifestyle covariates

For the main interaction analyses, the phenotypes of the main traits (Table S1) were adjusted for demographics, assessment centre, genotype measurement batch and population structure measured by the first 10 principal components (PCs) ^32^. Demographic variables included sex, year of birth, income, education, and Townsend deprivation index. The education variable was obtained following Okbay et al. (2016) ^33^, and the details is in Table S2. For each interaction analysis, the covariate in the interaction model was also used to adjust the phenotypes of the main trait, which was necessary to avoid any spurious interaction signals due to correlations between the main trait and the covariate ^27,34^. In addition to these key variables for the adjustment, other lifestyle covariates not in the model could be considered, depending on their relevance to the main trait.

When a lifestyle covariate was used as the second trait in a bivariate model (see Note S3), the phenotypes of the second trait were also adjusted for potential confounders including demographics, batch, centre and the first 10 PCs. In addition to these potential confounders, other lifestyle covariates were possibly considered, depending on their relevance to the second trait. The distribution of adjusted phenotypes of lifestyle covariates are shown in Figure S1.

For the diabetes-related metabolic traits in the main analyses, an additional quality control (QC) was applied to the adjusted phenotypes to exclude outliers that are outside the three standard deviations in either direction from the mean of the phenotypic data ^35^. The adjusted and QCed phenotypes were further transformed using a rank-based inverse normal function to satisfy the underlying assumption of the model, i.e. the normality assumption (see Figure S2). Note that these steps are essential to prevent spurious interaction signals ^27,34^. The number of individual records remained after these processes (adjustment, outlier QC and transformation) are listed in Table S3.

#### Statistical models

We used a multivariate reaction norm model (MRNM) that can estimate both GxE and residual-by-environment (RxE), simultaneously ^27^, where RxE indicates the non-genetic effects that are modulated by the lifestyle covariates. Data analysis were performed using MTG2 software ^36^. In the main analyses, there were four models, i.e. a null model without any interactions, model with GxE or RxE interaction only and full model jointly fitting GxE and RxE interactions. The maximum likelihoods from the four models were compared to test if there was significant interaction (see Figure S3). A significance p-value threshold was set at 5.21E-04 (=0.05 / 96) after Bonferroni correction to account for 96 tests in total. We declared a significance if the p-value was lower than the significance threshold and the sum of estimated variances of GxE and RxE (i.e., 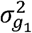 and/or 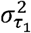) was non-negative to avoid estimated interaction effects out of the legitimate parameter space. The model description for MRNM can be found in Note S3.

#### Model Comparisons to detect interaction effects

Based on the model comparison using four different MRNMs (Note S3), five different interaction effects can be tested (Figure S3 & Table S4). The restricted maximum likelihood values obtained from the four models were used to assess their model fits ^27^. The significance of interaction effects was determined based on the p-values from likelihood ratio chi-squared tests comparing the full and reduced models. The five kinds of interaction effects are 1) overall interaction detected from the comparison between the null and full models, which includes both GxE and RxE interaction, noting that the overlapping section represents the collinearity between estimated GxE and RxE interactions ^27^; 2) GxE interaction detected from the comparison between the null and GxE only models, which is not corrected for the collinearity with RxE interaction; 3) RxE interaction detected from the comparison between the null and RxE only models, which is not corrected for the collinearity with GxE interaction; 4) orthogonal GxE interaction detected from the comparison between the RxE only and full models, which is corrected for the collinearity with RxE interaction; and 5) orthogonal RxE interaction detected from the comparison between GxE and full models, which is corrected for the collinearity with GxE interaction (Figure S3). It is noted that while overall interactions are important, it is of interest to disentangle between GxE and RxE interactions that are without collinearity, referred to as orthogonal GxE or RxE interaction (Figure S3).

#### Predicted phenotypes (risk) of metabolic traits

Based on the full model (see Note S3), the expected phenotypes for each individual, comprising of estimated additive genetic (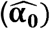), GxE interaction (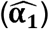) and RxE interaction effects 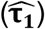), can be written as

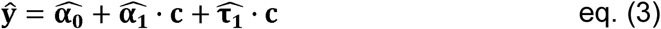

where ***ŷ*** is the predicted phenotypes, and **c** is the standardised lifestyle covariate that with a mean of 0 and a standard deviation of 1. To be consistent across traits, we used the full model to predict the phenotypes for all traits.

Furthermore, we derived the trajectory of the predicted phenotypes across different levels of lifestyle covariates in each of the 96 analyses (12 traits x 8 covariates). For this, individuals were divided into three groups, that is, the top, middle and bottom 20% groups according to the estimated interaction effects (the sum of GxE and RxE effects, i.e., 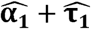). In addition, each of the three groups was further stratified into two groups, T2DM prospective cases and controls. Note that we restricted to use incident (i.e. prospective) cases only, according to ICD-10 information (code E11) to avoid any effects of T2DM status on metabolic risk (Note S1). We estimated the intercept and slope of a linear model regressing the predicted phenotypes on the covariate for each group.

The values of intercepts and slopes were averaged over the 68 analyses that showed significant signals for both GxE and RxE interactions. The averaged values of intercepts and slopes might represent the overall relationship between metabolic risk and lifestyle covariates. In this process, we considered making favourable and unfavourable directions consistent across the main traits and covariates, to facilitate a better interpretation in line with metabolic risks on T2DM. For example, the sign of HDL, physical activity and healthy diet values were switched when analysing phenotypes so that the direction of favourableness for these variables is consistent with other variables (glucose, HbA1c, TC, LDL, CRP, sodium, potassium, BMI, WHR, systolic BP, diastolic BP, Age, ALC, SMK). To test whether the difference between the groups with cases and controls is statistically significant, we did a paired t-test.

In addition to the comparison using 68 analyses with the significant overall interactions, we performed the same analyses using 17 analyses that showed significant signals for orthogonal GxE interaction and 56 analyses for orthogonal RxE interaction. The predicted phenotypes were grouped as same as the overall interaction according to the estimated of GxE or RxE interaction effects (i.e., 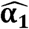 or 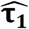.

#### Heritability

We calculated the heritability using the estimated genetic and residual variances from the null model (i.e. multivariate GREML) or interaction model (MRNM), which can be expressed as

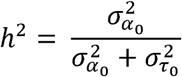

where 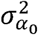 is the main genetic variance, and 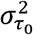 is the residual variance. This estimation assumes the environmental homogeneity of the study sample.

#### Causality Analysis

As complementary analyses, we used CAUSE software ^37^, which is a newly proposed Mendelian randomization method, to detect the causal effects of lifestyle covariates on T2DM metabolic traits. In causality analyses, the GWAS summary statistics of pruned SNPs were used to infer the causality and its significance, following the instruction of CAUSE. The same genotype and phenotype data as in the main interaction analyses were used in the causality analyses. We note that the phenotypes of the metabolic traits used as outcomes in causality analyses were required no to be adjusted for the covariate that used as the exposure (i.e. lifestyle covariate) because CAUSE tests the association of the first moment (mean), differed from MRNM that is to estimate the second moment (variance), across covariate values.

## Results

### Interactions

We tested if the genetic (GxE) and non-genetic effects (RxE) of 12 diabetes-related metabolic traits are modulated by eight lifestyle covariates (see Methods) and found that 68 out of 96 tests showed significant signals for the interactions when comparing the full and null models (Figure S4). The significant p-values were obtained from the likelihood ratio tests after Bonferroni correction. As shown in Figure S4, the genetic and non-genetic effects of glucose and HbA1c, which are highly associated with T2DM (Table S5), are shown to be significantly modulated by most of the 8 lifestyle factors. CRP, which is a well-known biomarker of inflammation, is significantly altered by all lifestyle covariates.

It is of interest to disentangle the modulated genetic effects from non-genetic effects, i.e. GxE effects orthogonal to RxE effects. When comparing the full model with RxE only model, there were 17 tests showing significant orthogonal GxE interactions after the Bonferroni correction (see Methods) (Figure 1 & Table S6). Specifically, the genetic effects of glucose were shown to be modulated by the level of physical activity (p-value=1.12E-04) and ALC (p-value=5.12E-05). Strong modulations of genetic effects by physical activity were observed for HbA1c and CRP (p-values < 4.66E-04). Although there was no evidence of orthogonal interaction between physical activity and BMI, other lifestyle factors significantly modulated the genetic effects of BMI (e.g. p-value = 1.86E-10 for ALC-BMI) (Figure 1).

**Figure 1.**
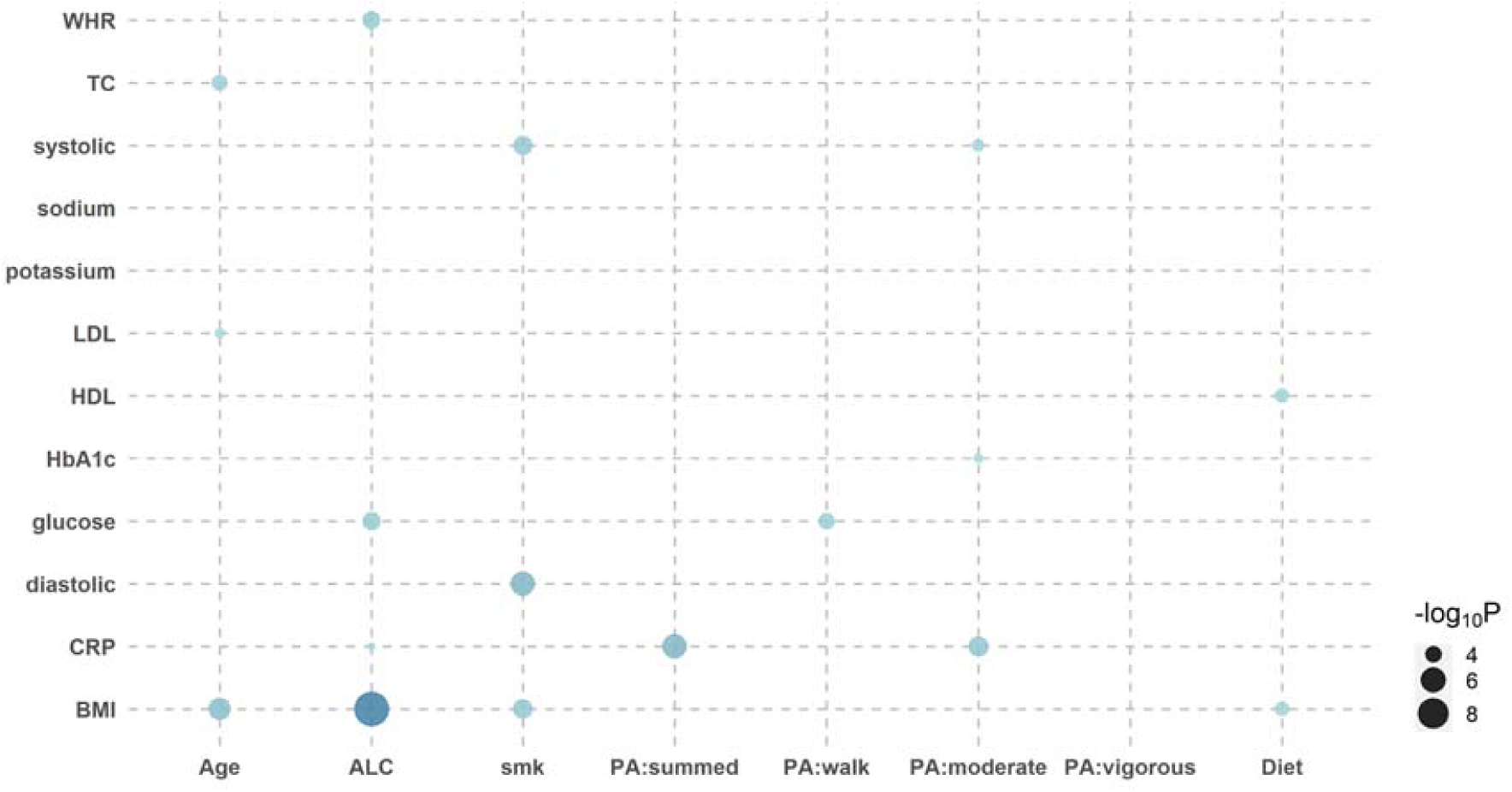
Bubble plot of p-values indicating significant orthogonal GxE interactions. There were 17 significant orthogonal GxE interactions when testing the genetic effects of 12 diabetes-related metabolic traits modulated by 8 covariates. Likelihood ratio tests were used to compare the full model with the RxE only model for each of 6 independent datasets, and p-values were meta-analysed using the Fisher’s method. The size of dot reflects its significance, the bigger the more significant. For example, the BMI-ALC pair is most significant (p-value= 1.86E-10), indicating that ALC significantly modulates genetic effects of BMI phenotypes. *WHR; waist-hip ratio, TC; Total cholesterol, systolic; systolic blood pressure, sodium; sodium in urine, potassium; potassium in urine, LDL; low density lipoprotein cholesterol, HDL; high density lipoprotein cholesterol, HbA1c; haemoglobinA1C, CRP; C reactive protein, BMI; body mass index, Age; Age at recruitment, ALC; alcohol intake frequency, smk; smoking status, PA:summed; a summed physical activity, PA:walk; physical activity walking, PA:moderate; physical activity moderate intensity, PA:vigorous; physical activity vigorous intensity, Diet; healthy dietary scores*,

We also compared the full model with GxE only model, to assess orthogonal RxE interaction effects, and found 56 significant signals out of 96 tests (Figure S5 & Supplementary File 1). The significance of RxE interaction was generally stronger than that of GxE interaction. For glucose and HbA1c, which are closely related to T2DM, most of the lifestyle factors had significant modulation effects, captured by orthogonal RxE interaction. It is remarkable that there were significant interaction signals for CRP that were consistently observed across all lifestyle factors with p-values ranging from 4.32E-96 to 5.51E-12. In the analyses of BMI, a strong risk factor of T2DM, it was shown that the non-genetic effects ^38^ of BMI were significantly modulated by ALC and physical activity.

A number of orthogonal RxE interactions found in this study can be supported by a causality analysis, using CAUSE software ^37^ (Figure S6). For example, CAUSE analysis showed significant causal effects of lifestyle factors on phenotypes for pairs of ALC-HbA1c, ALC-TC, ALC-HDL and Diet-Sodium, which also appeared to be significant for the orthogonal RxE interaction (Figure S5). In addition, the significant causal relationship of each pair of ALC-BMI, ALC-WHR and SMK-WHR can be partly explained by the orthogonal GxE interaction (Figure 1) or combined GxE and RxE interactions (Figure S2).

MRNM allows individually different responses to a lifestyle modification, which cannot be modelled in standard additive models. To demonstrate this property of MRNM, we plotted predicted phenotypes (see eq. (3) in Methods) against the standardised values of lifestyle factors for three groups (the top, middle and bottom 20%) stratified according to the magnitude of estimated GxE interaction (Figure S7). This shows that the expected phenotypes in responses to the modification of lifestyle factors can be different among individuals, and the slope of phenotypes is positive, zero or negative for the top, middle or bottom group, respectively (Figure S7). This demonstrates that individual genetic difference should be carefully considered in a lifestyle modification, i.e. an intervention of metabolic risk.

We further stratified each of the three groups into T2DM prospective cases and controls, hence six groups in total. The intercept and slope of regressing the predicted phenotypes on the standardised lifestyle measures were estimated for each of the six groups (e.g., Figure S8). We applied this approach to the 68 pairs with significant overall interactions (Figure 2) and calculated the mean and standard error of intercepts and slopes across all pairs, to assess if there was any significant difference between prospective cases and controls in each of the bottom, middle and top 20% groups (Figure 2). It was shown that the intercepts of prospective cases were significantly higher than those of controls in all three groups (paired t-test p-value=2.58E-07, 7.16E-10 and 2.78E-09), indicating that prospective cases were likely to have higher metabolic risk than controls. However, the slope of predicted risks was not significantly different between prospective cases and controls, showing that the change of metabolic risk in response to lifestyle modification is invariant across prospective cases and controls. Similar results were observed when considering GxE or RxE interaction only in that the intercepts were higher for prospective cases than controls (Figure S9 & S10). We note that when using the model of RxE interaction only, the slope was significantly steeper for prospective cases than controls in each of the three groups, suggesting that the metabolic risk of prospective cases is more sensitive to lifestyle modification, compared to controls (Figure S10), which was, however, not observed when using the model of GxE interaction only.

**Figure 2.**
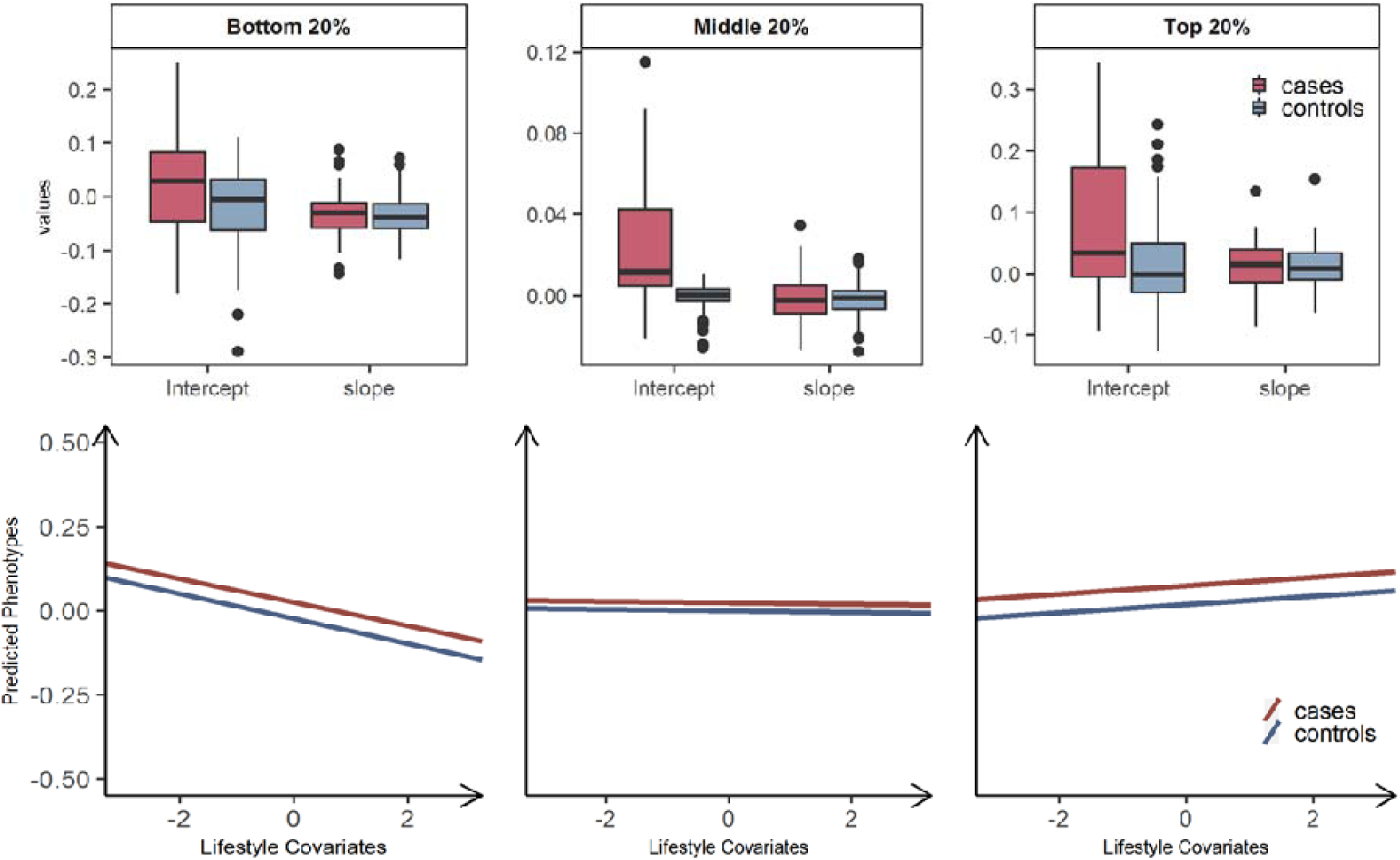
Plasticity of diabetes-related metabolic traits of 68 significant overall interactions in response to the level of lifestyle effects, by grouping T2DM prospective cases and controls. Individuals are stratified into three groups, the bottom, middle and top 20% groups according to estimated GxE and RxE interactions from the full model (i.e. 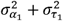). We further classified individuals into T2DM prospective cases (red) and controls (blue) in each of three groups, hence six groups in total. To compare the differences in trajectories between with T2DM cases and controls, a linear model regressing the phenotype of main trait on standardised lifestyle measures was used to estimate the intercept and slope for each of six groups. The values of intercepts and slopes were averaged over the 68 analyses that showed significant signals for both GxE and RxE interactions. The averaged values of intercepts and slopes represent the overall relationship between metabolic risk and lifestyle covariates, where we considered making favourable and unfavourable directions consistent across the main trait and covariates, to facilitate a better interpretation in line with metabolic risks on T2DM (see Methods). Box plot for each group was represented to show the differences in terms of the calculated intercepts and slopes, and the mean of values for intercepts and slopes were represented as regression lines. The mean and standard error of intercepts and slopes across the analyses of the 68 pairs with significant overall interactions were estimated to assess if there is any significant difference of the mean between cases and control in each of the three groups, the bottom, middle and top 20% groups. There were no significant differences between cases and controls in the mean of slopes in all three groups. The mean of intercepts is significantly different between cases and controls in all three groups (p-values= 2.58E-07, 7.16E-10 and 2.78E-09). The arrows on both axes in linear regression figures indicate an unfavourable direction in regard to T2DM health.

### Heritability

Heritability estimation can be biased if non-negligible interaction effects are not properly modelled. In the standard additive model (e.g. GREML), the variance attributed to un-modelled interactions can be partitioned as residual variance, which results in underestimated heritability, i.e. so-called still-missing heritability ^39^. This is evident for diabetes-related metabolic traits (Figure 3), for which the ratio of change in heritability for each trait was positive and not negligible. For example, the estimated heritability of CRP increased by 2.5% when significant interactions were considered appropriately. In the comparison of glucose and HbA1c, which are most relevant to T2DM, we observed more than 1.5% of heritability changes, and this supports the hypothesis that a fraction of still-missing heritability in metabolic traits is due to interaction effects being unaccounted for. The estimated variances for the main genetic and residual effects from GREML and MRNM were compared, showing that the residual variance was mostly overestimated (hence underestimated heritability) in the GREML (Figure S11).

**Figure 3.**
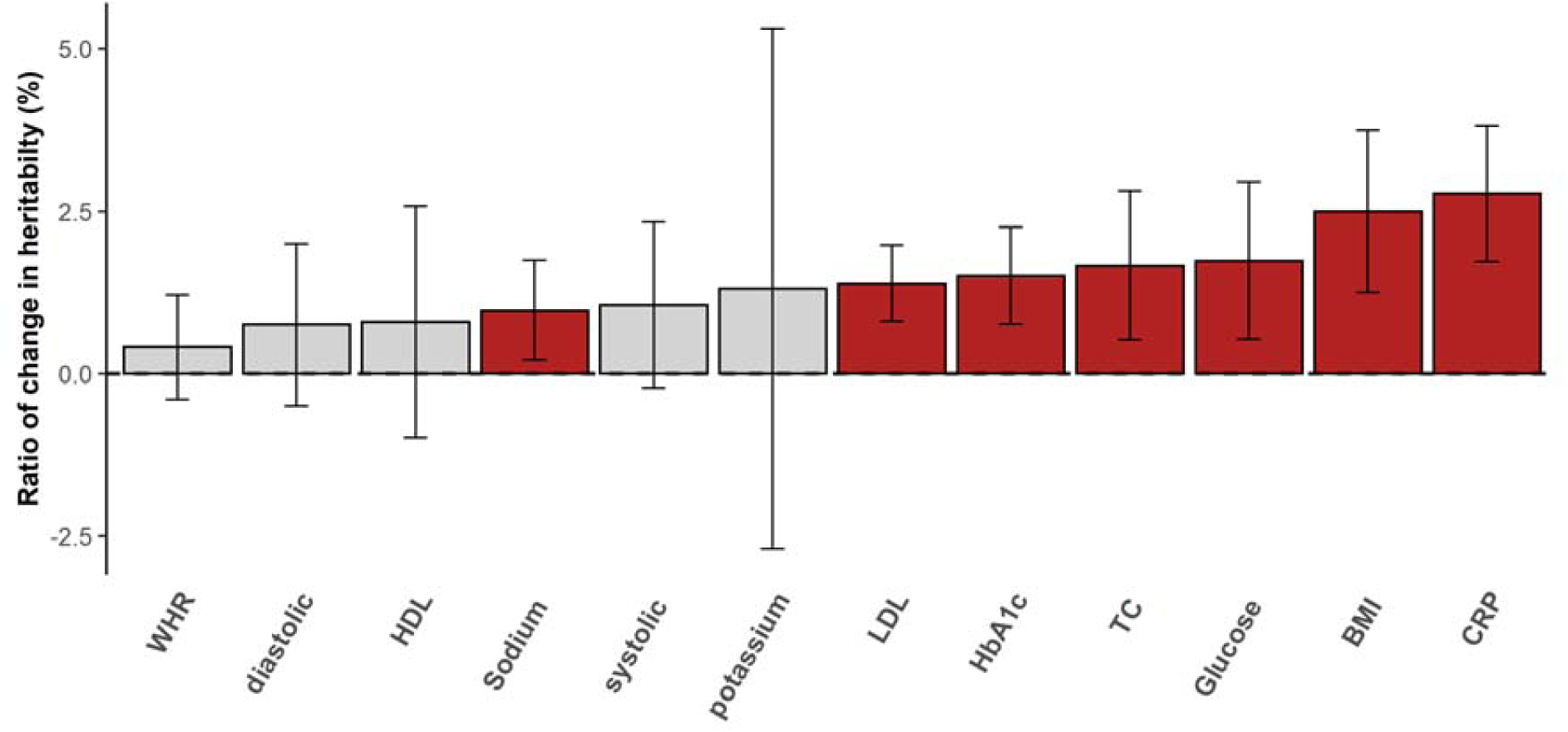
The ratio of change in SNP heritability. The differences between heritability estimates from the GREML and MRNM were represented as the ratio of change in heritability (%). Each bar indicates the differences in heritability, and the ratio was calculated as 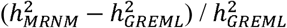 Where 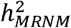 and 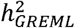 indicate the estimated heritability from MRNM and GREML, respectively. Therefore, the positive ratio denotes that estimate of MRNM is higher than that of GREML. The vertical line is the 95% confidence interval of the ratio averaged over the analyses of each trait with significant overall interaction. Traits with significant difference between MRNM and GREML were coloured as red. The interaction analyses of 68 pairs with significant overall interactions were used in the heritability comparisons. *WHR; waist-hip ratio, diastolic; diastolic blood pressure, HDL; high density lipoprotein cholesterol, sodium; sodium in urine, LDL; low density lipoprotein cholesterol, potassium; potassium in urine, TC; total cholesterol, HbA1c; haemoglobinA1C, BMI; body mass index, CRP; C reactive protein, systolic; systolic blood pressure*.

## Discussion

It is well established that phenotypic changes in diabetes-related metabolic traits such as glucose and HbA1c are associated with lifestyle modifications ^40-42^, which has motivated lifestyle interventions for the prevention and treatment of T2DM ^43-45^. However, it remains unknown if these interventions should be applied uniformly to everyone (i.e., a one-size-fits-all approach) or tailored to individual (i.e., precision health approach). In this study, we show that lifestyle modification can significantly alter the genetic and non-genetic effects of metabolic traits (i.e. GxE and RxE interactions), where the direction and magnitude of the alteration depend on individual differences in genetic information. This finding demonstrates that a more personalised approach is needed for T2DM intervention.

Previous studies have already indicated concerns about the inefficiency of the one-size-fits-all approach ^26,46-48^. These concerns can be overcome by accounting for individual genetic difference. For example, it is desirable to know how the diabetes-related metabolic risk in response to lifestyle modifications varies across individuals according to their genotypes, and this knowledge will allow a personalised lifestyle intervention to T2DM that can be tailored to each individual need.

Our finding for significant genome-wide GxE interactions across diabetes-related metabolic traits is novel and can be applied in such personalised lifestyle interventions. To our knowledge, no whole-genome GxE interactions have been reported for diabetes-related metabolic traits. Previously, we reported significant GxE and RxE interactions for some metabolic traits including BMI, BP, cholesterols and WHR, however, GxE interaction could not be disentangled from RxE because of a small sample size ^27,49^. In this study, we disentangled GxE interaction from RxE interaction using a large sample size for diabetes-related metabolic traits such as glucose, HbA1c and CRP that were not studied before. We also demonstrated the validity of the estimated RxE interactions, using CAUSE analyses ^37^. RxE interaction can be also explained by environment-by-environment interactions ^38^.

Unlike standard additive models, MRNM allows us to stratify samples into three groups (the top, middle and bottom 20%) according to estimated individual GxE or/and RxE interaction effects. The patterns of the expected phenotypes of the metabolic traits were clearly distinct between the three groups. This shows that the one-size-fits-all approach may not be the best strategy in a T2DM intervention. Importantly, the predicted metabolic risk was significantly higher in T2DM prospective cases than controls. Interestingly, the phenotypic plasticity of metabolic risk in response to lifestyle modification is significantly different between prospective cases and controls only when considering non-genetic effects of metabolic risk (i.e. using the model with RxE interaction only).

We found that 68 significant signals out of 96 tests were detected for overall interactions, indicating that GxE and RxE interactions play a significant role in the etiology of T2DM. To disentangle GxE from RxE, we adjusted the significance of GxE effects accounted for the collinearity with RxE and found 17 significant signals for orthogonal GxE interactions. Similarly, we found 56 significant signals for orthogonal RxE interactions. The smaller number of significant GxE interactions, compared to RxE interactions, is probably due to the fact that the power is smaller because of a large number of genetic variants (> 1M) were involved in the interaction term ^50^.

Although a causality analysis (CAUSE) was used to replicate some of our findings, we note that causality models (such as CAUSE or Mendelian randomization model) are different from MRNM in that they test associations among genetic instruments, exposure and outcome at the phenotypic level using a least square or similar methods. MRNM, instead, adjusts and removes the phenotypic association because its main interest is to estimate the heterogeneity of genetic and non-genetic variance across different lifestyle values. Therefore, MRNM is probably robust to the assumptions to be made in the causality models (e.g. horizontal pleiotropic effects that are removed from the adjustment in MRNM). Nonetheless, consistent results from these two very different models can increase the reliability of the findings.

In summary, the modulation of diabetes-related metabolic risk in response to the level of lifestyle covariates was clearly observed, and its direction and magnitude varied depending on individual genetic differences. Interestingly, such genetic phenotypic plasticity was invariant across T2DM prospective cases and controls although the overall metabolic genetic risk is significantly higher in T2DM prospective cases than controls. Our findings highlight the importance of individual genetic differences in the prevention and management of diabetes and suggest that the one-size-fits-all approach may not benefit all.

## Data Availability

The analyses conducted in this study were based on data accessed through the UK Biobank HTTP://www.ukbiobank.ac.uk

HTTP://www.ukbiobank.ac.uk

## Acknowledgments

We would like to thank all participants and staff of the UK Biobank for their valuable contributions. This research has been conducted using UK Biobank resource with application 14575, and the Research Ethics Committee (REC) approval number is 11/NW/0382. The work was supported by computational resources provided by the Australian Government through Gadi HPC under the National Computational Merit Allocation Scheme. We also thank UniSA Cancer Research Institute to support this research.

