## Supplementary documents for "Lifestyle modifies the diabetes-related metabolic risk, conditional on individual genetic differences"

**Index**

1. **Supplementary Note**

Note S1. phenotypic data and variables

Note S2. Calculation of the healthy dietary scores using the following variables in the UK Biobank.

Note S3. Statistical models

2. **Supplementary Table**

Table S1. Key variables information in the UK Biobank

Table S2. The details of converting the qualification levels to the education levels

Table S3. The number of individuals used in the main analyses, after QC, adjustment and rank-based inverse normal transformation

Table S4. Model comparison for detecting five kinds of interaction effects

Table S5. Linear regression coefficients between T2DM and 12 metabolic traits

Table S6. Estimates with significant signals for orthogonal GxE interaction from the meta-analysed results

3. **Supplementary Figure**

Figure S1. Histograms for the lifestyle covariate that showed their distribution of scaled only and pre-adjusted

Figure S2. Histograms for the T2DM traits that showed their distribution before and after applying RINT

Figure S3. The illustration of five different model comparisons

Figure S4. Bubble plot of p-values indicating significant overall interactions (i.e. GxE and/or RxE)

Figure S5. Bubble plot of p-values indicating significant orthogonal RxE interactions

Figure S6. Heatmap plot for the causality

Figure S7. Individuals phenotypic effect of six traits with the most significant overall interaction in response to the level of environmental covariates

Figure S8. Plasticity of six traits with the most significant overall interaction in response to the level of environmental effects, by grouping T2DM prospective cases and controls.

Figure S9. Plasticity of diabetes-related metabolic traits of 17 significant GxE interactions in response to the level of lifestyle effects, by grouping T2DM prospective cases and controls.

Figure S10. Plasticity of diabetes-related metabolic traits of 56 significant RxE interactions in response to the level of lifestyle effects, by grouping T2DM prospective cases and controls.

Figure S11. Variance differences between the bivariate GREML and MRNM analyses in genetic and non-genetic effects

### Supplementary Note

#### 1. **Phenotypic data and variables**

##### 1.1. Main traits

###### 1.1.1. Blood biochemistry

The biochemistry markers measured from blood samples of initial assessment visit participants were collected, e.g., glucose, Haemoglobin A1c, total cholesterol, LDL direct, HDL cholesterol, C-reactive protein. Glucose, of type continuous, was measured by hexokinase analysis from 429,568 participants. Haemoglobin A1c (HbA1c), of type continuous, was measured by HPLC analysis from 466,505 participants. Total cholesterol (TC), of type continuous, was measured by CHO-POD analysis from 469,590 participants. LDL direct (LDL), of type continuous, was measured by enzymatic protective selection analysis from 468,707 participants. HDL cholesterol (HDL), of type continuous, was measured by enzyme immunoinhibition analysis from 429,872 participants. C-reactive protein (CRP), of type continuous, was measured by immunoturbidimetric from 468,569 participants.

###### 1.1.1. Urine assays

The urinary biomarkers (e.g., sodium and potassium) was collected using ion-selective electrode method to measure the concentration in the stored urine sample. Sodium concentration in urine (sodium), of type continuous, was collected from 483,279 participants, and potassium concentration in urine (potassium), of type continuous, was collected from 483,290 participants.

###### 1.1.2. Physical measures

Various physical measurements were taken from the participants in UK Biobank, of which body mass index, waist-hip ratio, blood pressures were used in the analysis. *Body mass index* (BMI) was calculated by dividing the weight (kg) by the measures standing height (m) from 499,400 participants. *Waist-hip ratio* (WHR) was calculated from the waist circumference/Hip circumference(cm), where the waist circumference was measured from 500,345 participants, and hip circumference was from 500,286 participants. Both types of systolic and diastolic blood pressures were automatically measured two times with short term apart, and each blood pressure was obtained from the average of two records. *Diastolic blood pressure* (diastolic), of type integer, was collected from 472,392 participants, and *systolic blood pressure* (systolic), of type integer, was from 472,387 participants.

##### 1.2. Lifestyle covariates

Lifestyle covariates were used as the environmental variables in the GxE analysis to investigate if they modulate the genetic and non-genetic effects of T2DM metabolic traits. To estimate the genetic and non-genetic risk modulated by lifestyle covariates for the metabolic traits, eight lifestyle covariates were selected. All lifestyle covariates were obtained from UK Biobank participants from the touchscreen questionnaire during the first survey between 2006 and 2010. *Age at recruitment* (Age), of type integer, was collected from 502,505 participants based on the date of birth and their participant date. *The smoking Status* (SMK) was obtained by combining two datasets which are pack-years of smoking and Ever smoked. Since the pack-years of smoking were based on people who answered “yes” to Ever smoked question, participants who answered “No” were coded smoking status as zero. *Alcohol intake frequency* (ALC) was determined based on the question about “how often do you drink alcohol?” (Data-field: 1558), and all participants who responded “prefer not to answer” were excluded. *Physical Activity* (PA) variable in UK Biobank collected using international physical activity questionnaire short form (IPAQ-S) that categorised into three types of physical activity: 1) Walking; 2) Moderate; 3) Vigorous according to the intensity. All participants answered questions about “How many minutes per week of their engagement in the three categories physical activities”, and those are expressed in MET-minutes per week following the estimated MET-levels for each type of physical activities; Walking =3.3 METs, Moderate-intensity=4.0 METs and Vigorous-intensity= 8.0 METs. The continuous physical activity score quantified as MET-level * minutes of activity/day* days per week. In addition, a total physical activity score (summed) was calculated from the sum of three categories. *Healthy diet score* (Diet). Following the American Heart Association Guidelines ^1^ and Loes et al (2018) ^2^, the continuous dietary scores were calculated for fitting to multivariate reaction norm model as the lifestyle covariate. We defined four categories for the dietary patterns, which are fruit and vegetable intake, total fish intake, processed meat intake and red meat intake (see the details of data information in Note S2). Each of 4 dietary factors was given a score of 1 point, and the overall dietary score consist of a minimum 0 point and a maximum of 4 point for each participant. In summary, individuals with a score close to 4 can be represented as indicators of healthier diet habits than individuals with a score close to zero.

### 1.3. T2DM

Incident (prospective) cases of T2DM were identified by the ICD-10 (code E11), of who were excluded if the date E11 first reported is before the date of attending assessment centre. In addition, participants who were not cases in ICD-10 but if they were patients based on the self-reported information were excluded. Self-reported T2DM case-control status was determined based on information on diabetes diagnosed by doctor, excluding the patients who were aged under 35 at diagnosed diabetes and who had been prescribed insulin within a year (see the variables information in Table S1).

#### 2. **Calculation of the healthy dietary scores using the following variables in the UK Biobank.**

##### 2.1. Dietary variables in the UK Biobank

| **Variables** | **Data-Field** | **Questions** | **Data-coding** |
| --- | --- | --- | --- |
| **category 1. Fruit and Vegetables** | | | |
| Fresh fruit intake | 1309 | About how many pieces of FRESH fruit would you eat per day | Value Type: Integer (0~50),  -10: Less than one  -1: Do not Know  -3: Prefer not to answer |
| Cooked vegetable intake | 1289 | On average how many heaped tablespoons of COOKED or SALAD/RAW vegetables would you eat per DAY? | Value Type: Integer (0~50),  -10: Less than one  -1: Do not Know  -3: Prefer not to answer |
| Salad/Raw Vegetable intake | 1299 |  |  |
| **Category 2. Total fish Intake** | | | |
| Oily fish^1^ intake | 1329 | How often do you eat oily or non-oily fish? | 0: Never  1: Less than once a week  2: Once a week  3: 2-4 times a week  4: 5-6 times a week  5: Once or more daily  -1: Do not know  -3: prefer not to answer |
| Non-oily fish intake | 1339 |  |  |
| **Category 3. Processed meat** | | | |
| Processed meat intake | 1349 | How of often do you eat processed meats (e.g. bacon, ham, sausages, meat pies, kebabs, burgers, chicken nuggets)? | 0: Never  1: Less than once a week  2: Once a week  3: 2-4 times a week  4: 5-6 times a week  5: Once or more daily  -1: Do not know  -3: prefer not to answer |
| **Category 4. Red meat** | | | |
| Beef intake | 103020 | How many servings of beef or Lamb/mutton or Pork did you have? | 0: Never  1: Less than once a week  2: Once a week  3: 2-4 times a week  4: 5-6 times a week  5: Once or more daily  -1: Do not know  -3: prefer not to answer |
| Lamb/mutton intake | 103040 |  |  |
| Pork intake | 103030 |  |  |

^1^Oily fish: Salmon Anchovies, Trout Swordfish, Mackerel Bloater, Herring Cacha, Sardines Carp, Pilchards Hilsa, Kipper Jack fish, Eel Katla, Whitebait Orange roughy, Tuna (fresh only) Pangas, Sprats

##### 2.2. Total fruit and vegetables intake

For fresh fruit intake, a piece of fruit intake assumed as a serving. Total vegetable intake was obtained by combining COOKED and SALAD/RAW vegetable intake variables, and each of records coded as tablespoons were calculated as per servings by dividing into 3 (i.e. one serving was considered as 3 Tablespoons). The amount of total fruit and vegetable intake per day was obtained by combining the amount of total fruit and vegetable intake. To calculate healthy dietary scores, the variables of individuals who had 4.5 or more servings a day were converted to 1, and the variables of remains were converted to 0.

##### 2.3. Total fish intake

Before combining the variables, each of the variables for fish intake was recorded based on the number of times taken a week as follows:

|  | UK Biobank Data-coding | Recoded in the basis of a week |
| --- | --- | --- |
| Never | 0 | 0 |
| Less than once a week | 1 | 0.5 |
| Once a week | 2 | 1 |
| 2-4 times a week | 3 | 3 |
| 5-6 times a week | 4 | 5.5 |
| Once or more daily | 5 | 7 |
| Do not know | -1 | NA |
| Prefer not to answer | -3 | NA |

The information about how often the participants eat total fish a week obtained from combining the recoded data of Oily and Non-oily fish intake. To calculate healthy dietary scores, the records of individuals who had 2 or more times per week were converted to 1, and the variables of remains were converted to 0.

##### 2.4. Total processed meat intake

The information about how often the participants eat total processed meat a week was obtained from the recoded data in the basis of a week. To calculate healthy dietary scores, the records of individuals who had 2 or fewer times per week were converted to 1, and the variables of remains were converted to 0.

##### 2.5. total red meat intake

The information about how often the participants eat total red meat a week was obtained from combining the recoded data in the basis of a week beef, lamb/mutton and pork intake data. To calculate healthy dietary scores, the records of individuals who had 5 or fewer times per week were converted to 1, and the variables of remains were converted to 0.

##### 2.6. Scoring for healthy diet habits

Following the above procedures, which are converting the diet pattern records of each individual into the score to determine the favourable/unfavourable (healthy/unhealthy) dietary patterns, we generated the continuous healthy diet scores. The continuous dietary score calculated for fitting to the reaction norm model based on the procedures of collecting binary healthy dietary pattern which has been used in the previous studies. Each of 4 dietary factors was given a score of 1 point, and the overall dietary score consist of a minimum 0 point and a maximum of 4 points for each participant. In summary, individuals with a score close to 4 can be represented as indicators of healthier diet habits than individuals with a score close to zero.

#### 3. **Statistical models**

##### 3.1. Linear Mixed model

A univariate linear mixed model (LMM) consists of fixed effects and random effects. Assuming that every individual has a single measured phenotype (i.e. no missing and no repeated measures), the model can be written as

|  | $\mathbf{y}=\boldsymbol{\mu}+\mathbf{g}+\mathbf{e}$ | eq. (1) |
| --- | --- | --- |

where $\mathbf{y}$ is a vector containing pre-adjusted phenotypes of n individuals, $\boldsymbol{\mu}$ indicates a vector after adjusting for the fixed effects such as demographics (e.g. sex, year of birth), $\mathbf{g}$ indicates a vector of random genetic effects, $\mathbf{e}$ is a vector of random residual effects. The variance of phenotypes for the trait can be expressed as

|  | $var\left( \mathbf{y} \right)\boldsymbol{=}\mathbf{A}\sigma_{\mathbf{g}}^{2}\boldsymbol{+}\mathbf{I}\sigma_{\mathbf{e}}^{2}$ | eq. (2) |
| --- | --- | --- |

where $\mathbf{A}$ represents the $n x n$ genomic relationship matrix between the individuals based on whole-genome SNP information, and $\mathbf{I}$ is the $n x n$ identity matrix. The genomic relationship matrix $\mathbf{A}$ can be obtained by calculating$\mathbf{W}\mathbf{W}^{'}/m$, where $\mathbf{W}$ is a column-standardised n x m matrix containing the genotype information of n individuals and m SNPs. The terms $\sigma_{\mathbf{g}}^{2}$ and $\sigma_{\mathbf{e}}^{2}$ are the genetic and residual variance for $\mathbf{y}$, respectively.

##### 3.2. Univariate Reaction Norm Model (RNM)

###### 3.2.1. GxE model

A reaction norm can be defined as the trajectory of phenotypic change of a single genotype across an environmental gradient, i.e. genotype-environment interaction. It can be termed as Genotype x Environment (GxE) interaction due to the lifestyle covariates which are commonly used as the environmental gradients. Following Ni, van der Werf ^3^, the RNM with a first order random regression can be written as

$$\mathbf{y}=\boldsymbol{\mu}+\mathbf{g}+\mathbf{e}=\boldsymbol{\mu}+\boldsymbol{\alpha}_{\mathbf{0}}+\boldsymbol{\alpha}_{\mathbf{1}}\boldsymbol{\cdot}\mathbf{c}+\mathbf{e}$$

where $\mathbf{g}$ can be decomposed into genetic random regression coefficients ($\boldsymbol{\alpha}_{\boldsymbol{0}},\boldsymbol{\alpha}_{\mathbf{1}})$ to account for GxE interaction and $\mathbf{c}$ is the standardised lifestyle covariate that modulates the phenotypic change used as the environmental gradient. The variance and covariance matrix of random regression coefficients for genetic effects ($\mathbf{K}_{\mathbf{y}}$) can be represented as

$$\mathbf{K}_{\mathbf{y}}=cov\left( \boldsymbol{\alpha}_{\boldsymbol{0}},\boldsymbol{\alpha}_{1} \right)=\left[ \begin{matrix} var(\boldsymbol{\alpha}_{\mathbf{0}}) & cov(\boldsymbol{\alpha}_{\mathbf{0}},\boldsymbol{\alpha}_{\mathbf{1}}) \\ cov(\boldsymbol{\alpha}_{\mathbf{1}},\boldsymbol{\alpha}_{\mathbf{0}}) & var(\boldsymbol{\alpha}_{\mathbf{1}}) \end{matrix} \right]$$

where $\boldsymbol{\alpha}_{\mathbf{0}}$ or $\boldsymbol{\alpha}_{\mathbf{1}}$ is an $n$ vector of the zero or first order of random regression coefficients for the covariate values.

The k x 2 matrix of $\boldsymbol{\phi}$, which is the zero and first order polynomials of$k$ covariate values, can be represented as

$\boldsymbol{\phi}=\left[ \begin{matrix} c_{1}^{0} & c_{1}^{1} \\ \vdots& \vdots\\ c_{k}^{0} & c_{k}^{1} \end{matrix} \right]=\left[ \begin{matrix} 1 & c_{1}^{1} \\ \vdots& \vdots\\ 1 & c_{k}^{1} \end{matrix} \right]$.

Assuming that each individual has a unique covariate value ($n$=$k$), the variance and covariance matrix of additive genetic effects (var ($\mathbf{g}$)) across covariate values can be obtained as

$$\mathrm{var}\left( \mathbf{g} \right)= \left[ \begin{matrix} \sigma_{g_{1}}^{2} & \cdots& \sigma_{g_{1}g_{k}} \\ \vdots& \ddots& \vdots\\ \sigma_{g_{k},g_{1}} & \cdots& \sigma_{g_{k}}^{2} \end{matrix} \right]=\boldsymbol{\phi}\mathbf{K}_{\mathbf{y}}\boldsymbol{\phi}^{\mathbf{'}}$$

where $\sigma_{g_{p}}^{2}$ indicates the genetic variance for the $p$ ($p$ =1,…, $k$) th covariate value,$\sigma_{g_{p}g_{q}}$ is the genetic covariance between the individuals at the $p$ and $q$ th covariate values and $\boldsymbol{\phi'}$ indicates the transposed polynomial matrix of $\boldsymbol{\phi}$, which is the 2 x k matrix.

Therefore, the variance-covariance matrix of observed phenotypes reflecting GxE interaction can be extended from eq. (2) as

$$\mathrm{var}\left( \mathbf{y} \right)=\boldsymbol{A\circ}\left( \boldsymbol{\phi}\mathbf{K}_{\mathbf{y}}\boldsymbol{\phi}^{\mathbf{'}} \right)+\mathbf{I}\sigma_{\mathbf{e}}^{2}$$

where Hadamard product, i.e., $\boldsymbol{A\circ}\left( \boldsymbol{\phi}\mathbf{K}_{\mathbf{y}}\boldsymbol{\phi}^{\mathbf{'}} \right)$, was used to reflect the genomic relationship between the individuals.

###### 3.2.2. RxE model

The described RNM above is estimating the genetic heterogeneity to detect GxE interaction, it can be modified to a model with a non-genetic heterogeneity in the same context as the GxE interaction was reflected. The non-genetic effects, which are the remaining random effects except for genetic effects (i.e. residuals), are affected by the environmental gradients, and it causes the phenotypic modulation. This can be termed as Residual x Environment (RxE) interaction as the GxE interaction has been defined. The RNM with a first order random regression can be written as

$$\mathbf{y}=\boldsymbol{\mu}+\mathbf{g}\boldsymbol{+}\mathbf{e}\boldsymbol{=}\mathbf{b}+\mathbf{g}+\boldsymbol{\tau}_{\mathbf{0}}+\boldsymbol{\tau}_{\mathbf{1}}\boldsymbol{\cdot}\mathbf{c}$$

where **e** can be decomposed with residual random regression coefficients ($\boldsymbol{\tau}_{\boldsymbol{0}},\boldsymbol{\tau}_{\boldsymbol{1}})$ to account for RxE interaction. The variance and covariance matrix of random regression coefficients for residuals effects ($\mathbf{M}_{\mathbf{y}}$) can be written as

$$\mathbf{M}_{\mathbf{y}}=cov\left( \boldsymbol{\tau}_{\mathbf{0}},\boldsymbol{\tau}_{1} \right)=\left[ \begin{matrix} var(\boldsymbol{\tau}_{\boldsymbol{0}}) & cov(\boldsymbol{\tau}_{\boldsymbol{0}},\boldsymbol{\tau}_{\boldsymbol{1}}) \\ cov(\boldsymbol{\tau}_{\boldsymbol{1}},\boldsymbol{\tau}_{\boldsymbol{0}}) & var(\boldsymbol{\tau}_{\boldsymbol{1}}) \end{matrix} \right]$$

where $\boldsymbol{\tau}_{\boldsymbol{0}}$or $\boldsymbol{\tau}_{\boldsymbol{1}}$ indicate an n vector of the zero or first order of random regression coefficients for the covariate values. Assuming that each individual has a unique covariate value (n=k), the variance and covariance matrix of residual effects (var ($\mathbf{e}$)) can be obtained as

$$\mathrm{var}\left( \mathbf{e} \right)= \left[ \begin{matrix} \sigma_{e_{1}}^{2} & \cdots& \sigma_{e_{1}e_{k}} \\ \vdots& \ddots& \vdots\\ \sigma_{e_{k},e_{1}} & \cdots& \sigma_{e_{k}}^{2} \end{matrix} \right]=\boldsymbol{\phi}\mathbf{M}_{\mathbf{y}}\boldsymbol{\phi}^{\mathbf{'}}$$

where $\sigma_{e_{p}}^{2}$ indicates the residual variance for the $p$ th covariate value and $\sigma_{e_{p},e_{q}}$ is the residual covariance between the individuals at the $p$ and $q$ th covariate values.

Therefore, the variance-covariance matrix of observed phenotypes reflecting RxE interactions can be extended from eq. (2) as

$$\mathrm{var}\left( \mathbf{y} \right)=\mathbf{A}\sigma_{\mathbf{g}}^{2}\boldsymbol{+}\boldsymbol{I\circ(\phi}\mathbf{M}_{\mathbf{y}}\boldsymbol{\phi}^{\mathbf{'}})$$

###### 3.2.3. Full model

The full model can jointly capture both genetic and residual effects that are modulated by the different lifestyle covariate values by estimating heterogeneity for both effects simultaneously. This model can be expressed as

$$\mathbf{y}=\boldsymbol{\mu}+\mathbf{g}+\mathbf{e}=\boldsymbol{\mu}+\boldsymbol{\alpha}_{\mathbf{0}}+\boldsymbol{\alpha}_{\mathbf{1}}\boldsymbol{\cdot}\mathbf{c}\boldsymbol{+}\boldsymbol{\tau}_{\mathbf{0}}+\boldsymbol{\tau}_{\mathbf{1}}\boldsymbol{\cdot}\mathbf{c}\boldsymbol{.}$$

The variance and covariance matrix of the trait phenotypes (without missing values and repeated measures) which are reflecting both interactions can be extended from eq. (2) as

$$\mathrm{var}\left( \mathbf{y} \right)=\mathbf{A} var\left( \mathbf{g} \right)+ \mathbf{I}var\left( \mathbf{e} \right)$$

$$=\mathbf{A} \circ(\boldsymbol{\phi}\mathbf{K}_{\mathbf{y}}\boldsymbol{\phi}^{\mathbf{'}})+\boldsymbol{I\circ}(\boldsymbol{\phi}\mathbf{M}_{\mathbf{y}}\boldsymbol{\phi}^{\mathbf{'}})$$

##### 3.3. Multivariate RNM (MRNM) accounting for heterogeneous genetic and residual variances

Following Ni, van der Werf ^3^, it has been confirmed the finding that any correlation between the main traits and covariates can cause spurious interaction signals. MRNM can control such spurious signals by explicitly modelling genetic and residual correlations between the main phenotypes and covariates. To account their correlations, the covariate is also modelled of which having fixed and random effects in the same context as the main trait is. The model equation, which reflects both the main trait and covariate simultaneously to estimate unbiased interactions, can be represented as

$$\mathbf{y}=\boldsymbol{\mu}+\mathbf{g}+\mathbf{e}=\boldsymbol{\mu}+\boldsymbol{\alpha}_{\mathbf{0}}+\boldsymbol{\alpha}_{\mathbf{1}}\boldsymbol{\cdot}\mathbf{c}\boldsymbol{+}\boldsymbol{\tau}_{\mathbf{0}}+\boldsymbol{\tau}_{\mathbf{1}}\boldsymbol{\cdot}\mathbf{c}$$

$$\mathbf{c}^{\boldsymbol{*}}=\boldsymbol{\mu}+\boldsymbol{\gamma}+\boldsymbol{\varepsilon}$$

where $\mathbf{c}$ is an n vector of the covariate phenotypes for individuals, $\boldsymbol{\gamma}$ and $\boldsymbol{\varepsilon}$ are the random genetic and residual effects, respectively. Here, the covariate fitted as the second trait in the model was pre-adjusted for the fixed effects, denoted as $\mathbf{c}^{\boldsymbol{*}}$. The covariance between the genetic effects of covariate ($\boldsymbol{\gamma}$) and the genetic random regression coefficients of the main phenotypes ($\boldsymbol{\alpha}_{\boldsymbol{0}}$ and $\boldsymbol{\alpha}_{\boldsymbol{1}}$) can be expressed as

$\mathbf{k}_{\mathbf{y,}\mathbf{c}^{\boldsymbol{*}}}\boldsymbol{=}\left[ \begin{matrix} cov(\boldsymbol{\alpha}_{\mathbf{0}}\boldsymbol{,\gamma}) \\ cov(\boldsymbol{\alpha}_{\mathbf{1}}\boldsymbol{,\gamma}) \end{matrix} \right]$.

In a similar manner, the covariance between the residual effects of ($\boldsymbol{\varepsilon}$) and the residual random regression coefficients of the main phenotypes ($\boldsymbol{\tau}_{\boldsymbol{0}}$ and $\boldsymbol{\tau}_{\boldsymbol{1}}$) can be expressed as

$\mathbf{M}_{\mathbf{y}\boldsymbol{,}\mathbf{c}^{\boldsymbol{*}}}\boldsymbol{=}\left[ \begin{matrix} cov(\boldsymbol{\tau}_{\mathbf{0}}\boldsymbol{,\varepsilon}) \\ cov(\boldsymbol{\tau}_{\mathbf{1}}\boldsymbol{,\varepsilon}) \end{matrix} \right]$.

The variance and covariance matrix of genetic effects considering the main phenotypes and covariate jointly in the MRNM can be represented as

$$var\left( \mathbf{g},\boldsymbol{\gamma} \right)=\left[ \begin{matrix} var(\mathbf{g}) & cov(\mathbf{g},\boldsymbol{\gamma}) \\ cov(\boldsymbol{\gamma},\mathbf{g}) & var(\boldsymbol{\gamma}) \end{matrix} \right]=\left[ \begin{matrix} \boldsymbol{\phi}\mathbf{k}_{\mathbf{y}}\boldsymbol{\phi}^{'} & \boldsymbol{\phi}\mathbf{k}_{\mathbf{y,}\mathbf{c}^{\boldsymbol{*}}} \\ \mathbf{k}_{\mathbf{y,}\mathbf{c}^{\boldsymbol{*}}}^{\boldsymbol{'}}\boldsymbol{\phi'} & var(\boldsymbol{\gamma}) \end{matrix} \right]= \left[ \begin{matrix} \left( \begin{matrix} \begin{matrix} \sigma_{g_{1}}^{2} \\ \vdots\\ \sigma_{g_{k},g_{1}} \end{matrix} & \begin{matrix} \cdots\\ \ddots\\ \cdots\end{matrix} & \begin{matrix} \sigma_{g_{1},g_{k}} \\ \vdots\\ \sigma_{g_{k}}^{2} \end{matrix} \end{matrix} \right) & \left( \begin{matrix} \sigma_{g_{1},\gamma} \\ \vdots\\ \sigma_{g_{k},\gamma} \end{matrix} \right) \\ \left( \begin{matrix} \sigma_{\gamma,g_{1}} & \cdots& \sigma_{\gamma,g_{k}} \end{matrix} \right) & \sigma_{\gamma}^{2} \end{matrix} \right]$$

where $\sigma_{g_{p},\gamma}$ is the covariance between the genetic effects of the main phenotypes and covariate at the $p$ th covariate level , $\sigma_{\gamma,g_{p}}$is the transposed form of the $\sigma_{g_{p},\gamma}$, and $\sigma_{\gamma}^{2}$ is the genetic variance of $\mathbf{c}^{\mathbf{*}}$.

The variance and covariance matrix of residual effects considering the main phenotypes and covariate jointly in the MRNM can be represented as

$$var\left( \mathbf{e},\boldsymbol{\varepsilon} \right)=\left[ \begin{matrix} var(\mathbf{e}) & cov(\mathbf{e},\boldsymbol{\varepsilon}) \\ cov(\boldsymbol{\varepsilon},\mathbf{e}) & var(\boldsymbol{\varepsilon}) \end{matrix} \right]=\left[ \begin{matrix} \boldsymbol{\phi}\mathbf{M}_{\mathbf{y}}\boldsymbol{\phi}^{'} & \boldsymbol{\phi}\mathbf{M}_{\mathbf{y,c}} \\ \mathbf{M}_{\mathbf{y,c}}^{\boldsymbol{'}}\boldsymbol{\phi'} & var(\boldsymbol{\varepsilon}) \end{matrix} \right]=\left[ \begin{matrix} \left( \begin{matrix} \begin{matrix} \sigma_{e_{1}}^{2} \\ \vdots\\ \sigma_{e_{k},e_{1}} \end{matrix} & \begin{matrix} \cdots\\ \ddots\\ \cdots\end{matrix} & \begin{matrix} \sigma_{e_{1},e_{k}} \\ \vdots\\ \sigma_{e_{k}}^{2} \end{matrix} \end{matrix} \right) & \left( \begin{matrix} \sigma_{e_{1},\varepsilon} \\ \vdots\\ \sigma_{e_{k},\varepsilon} \end{matrix} \right) \\ \left( \begin{matrix} \sigma_{e_{1},\varepsilon} & \cdots& \sigma_{e_{k},\varepsilon} \end{matrix} \right) & \sigma_{\varepsilon}^{2} \end{matrix} \right]$$

where $\sigma_{e_{p},\varepsilon}$ is the covariance between the genetic effects of the main phenotypes and covariate at the $p$ th covariate level, and $\sigma_{\varepsilon,e_{p}}$ is the transposed form of the $\sigma_{e_{p},\varepsilon}$, and $\sigma_{\varepsilon}^{2}$ is the residual variance of $\mathbf{c}^{\mathbf{*}}$.

According to the information for the components described above, the variance-covariance matrix of the main trait (**y**) and covariate ($\mathbf{c}^{\mathbf{*}}$), jointly modelled in MRNM, can be written as

$$\mathrm{cov}\left( \mathbf{y},\mathbf{c}^{\boldsymbol{*}} \right)= \left[ \begin{matrix} var\left( \mathbf{y} \right) & cov\left( \mathbf{y},\mathbf{c}^{\mathbf{*}} \right) \\ cov\left( \mathbf{c}^{\mathbf{*}},\mathbf{y} \right) & var\left( \mathbf{c}^{\mathbf{*}} \right) \end{matrix} \right]$$

$$= \left[ \begin{matrix} \mathbf{A} \circ(\boldsymbol{\phi}\mathbf{K}_{\mathbf{y}}\boldsymbol{\phi}^{\mathbf{'}})+\boldsymbol{I\circ}(\boldsymbol{\phi}\mathbf{M}_{\mathbf{y}}\boldsymbol{\phi}^{\mathbf{'}}) & \boldsymbol{\phi}\mathbf{k}_{\mathbf{y,}\mathbf{c}^{\boldsymbol{*}}}\boldsymbol{+\phi}\mathbf{M}_{\mathbf{y,c}} \\ \mathbf{k}_{\mathbf{y,}\mathbf{c}^{\boldsymbol{*}}}^{\boldsymbol{'}} \boldsymbol{\phi}^{\mathbf{'}}\mathbf{+}\mathbf{M}_{\mathbf{y,c}}^{\boldsymbol{'}}\boldsymbol{\phi'} & \mathbf{A}\sigma_{\gamma}^{2}+\mathbf{I}\sigma_{\varepsilon}^{2} \end{matrix} \right]$$

where the first column of the first row is the same as the URNM full model described above.

### Supplementary Tables

#### **Table S1. Key variables information in the UK Biobank**

| **Variables** | **Data-Field** | **Details** | **Value type** |
| --- | --- | --- | --- |
| **Demographic variables** | | | |
| Sex | 31 | Sex of participants | Categorical^1^ (Single, 2) |
| UK Biobank assessment centre | 54 | The UK Biobank assessment centre at which participant consented | Categorical (single, 27) |
| Townsend deprivation index | 189 | Townsend deprivation index calculated immediately prior to participant joining UK Biobank | Continuous^2^ (-6.26 – 11) |
| Genotype measurement Batch | 22000 | - | Categorical (single, 108) |
| Year of Birth | 34 | Year of birth of participant (years) | Integral^3^ (1934-1971) |
| Average total household Income | 738 | “what is the average total income before tax received by your HOUSEHOLD?” | Categorical (single, 7) |
| Education (Qualification)^4^ | 6138 | Which of the following qualifications do you have? | Categorial (multiple, 8) |
| Age | 21022 | Age of the participant on the day they attended an initial assessment centre (years) | Integral (37-73) |
| **Lifestyle Covariates** | | | |
| Smoking status | 20160 | “Ever smoked” | Categorical (single, 2) |
|  | 20161 | “Pack years of smoking” | Continuous (0-336) |
| Alcohol Intake frequency | 1558 | “About how often do you drink alcohol?” | Categorical (single, 7) |
| Physical Activity: walking | 22037 | Metabolic Equivalent Task (MET) minutes per week for walking (minutes/week) | Continuous (0-4158) |
| Physical Activity: moderate | 22038 | Metabolic Equivalent Task (MET) minutes per week for moderate activity (minutes/week) | Continuous (0-5040) |
| Physical Activity: vigorous | 22039 | Metabolic Equivalent Task (MET) minutes per week for vigorous activity (minutes/week) | Continuous (0-10080) |
| Physical Activity: summed | 22040 | Total Metabolic Equivalent Task (MET) minutes per week for all activity including walking, moderate and vigorous activity (minutes/week) | Continuous (0-19278) |
| Healthy diet scores | *See Note S2* | | |
| **Diabetes-related metabolic traits** | | | |
| Body Mass Index (BMI) | 21001 | Body composition estimation by impedance measurement (Kg/m^2^) | Continuous (12.12-74.68) |
| Waist-Hip Ratio | 49 | Hip circumference (cm) | Continuous (20 - 197) |
|  | 48 | Waist Circumference (cm) | Continuous (30 - 195) |
| Systolic Blood Pressure | 4080 | Blood pressure, automated reading, systolic (mmHg) | Integral, multiple (56-268) |
| Diastolic Blood Pressure | 4079 | Blood pressure, automated reading, diastolic (mmHg) | Integral, multiple (30-148) |
| Glucose | 30740 | Measured by hexokinase analysis on a Beckman Coulter AU5800 (mmol/L) | Continuous (0.99-36.81) |
| Glycated haemoglobin (HbA1c) | 30750 | Measured by HPLC analysis on a Bio-Rad VARIANT ll Turbo (mmol/L) | Continuous (15-515.2) |
| Potassium in Urine | 30520 | Measured by ISE (ion selective electrode), Beckman Coulter AU5400 (millimole/L) | Continuous (2.4-200) |
| Sodium in Urine | 30530 | Measured by ISE (ion selective electrode), Beckman Coulter AU5400 (millimole/L) | Continuous (10-380.7) |
| LDL direct | 30780 | Measured by enzymatic protective selection, Beckman Coulter AU5800 (mmol/L) | Continuous (0.27-9.8) |
| HDL-cholesterol | 30760 | Measured by enzyme immune inhibition, Beckman Coulter AU5800 (mmol/L) | Continuous (0.22-4.4) |
| Total cholesterol | 30690 | Measured by CHO-POD, Beckman Coulter AU5800 (mmol/L) | Continuous (0.6-15.46) |
| C-reactive protein | 30710 | Measured by immunoturbidimetric, Beckman Coulter AU5800 (mg/L) | Continuous (0.08-79.96) |
| **T2DM case-control status** |  |  |  |
| Date of attending assessment centre | 53 | Date that participant attended a UK Biobank assessment centre | 2006/03/13-2010/10/01 |
| Source of report of E11 | 130709 | Source of the first code mapped to 3-character ICD10 E11, corresponds to “non-insulin-dependent diabetes mellitus”. |  |
| Date E11 first reported | 130708 | Date of the first occurrence of any code mapped to 3-character ICD10 E11 |  |
| Age diabetes diagnosed | 2976 | “What was your age when the diabetes was first diagnosed?” (years) | Integral (1-70) |
| Diabetes diagnosed by doctor | 2443 | “Has a doctor ever told you that you have diabetes?” | Categorial (single) |
| Prescribed insulin within a year | 2986 | “Did you start insulin within one year of your diagnosis of diabetes?” | Categorial (single) |

Categorical^1^ (type, the number of categories)

Continuous^2^ (minimum value - maximum value)

Integral^3^ (minimum value - maximum value)

Education (Qualification)^4^: See Table S2 for the details how the qualification levels for each individual were converted to the education levels.

#### **Table S2. The details of converting the qualification levels to the education levels.**

| UKBB (Data-Field 6138) | Coded in UKBB | ISCED mapping^1^ | Converted to Education level (years) |
| --- | --- | --- | --- |
| College or university degree | 1 | ISCED 5 | 20 |
| A levels/As levels or equivalent | 2 | ISCED 3 | 13 |
| O levels/GCSEs or equivalent | 3 | ISCED 2 | 10 |
| CSEs or equivalent | 4 | ISCED 2 | 10 |
| NVQ or HND or HNC or equivalent | 5 | ISCED 5 | 19 |
| Other professional qualifications e.g. nursing, teaching | 6 | ISCED 4 | 15 |
| None of the above | -7 | ISCED 1 | 7 |
| Prefer not to answer | -3 | NA | NA |

**^1^**ISCED mapping: International Standard Classification of Education

The education variable was collected based on the qualification questionnaire “which of the qualification do participants have?”. We followed the Okbay et al. (2016) ^4^ for converting the qualification information to the education levels for each individual. From the multiple responses, the highest education year, which is converted based on the qualification information, was selected for the education level of each participant.

#### **Table S3. The number of individuals used in the main analyses, after QC, adjustment and rank-based inverse normal transformation.**

| **The number of individuals for diabetes-related metabolic traits** | | | | | | | | |
| --- | --- | --- | --- | --- | --- | --- | --- | --- |
|  | **Age** | **ALC** | **SMK** | **PA: walking** | **PA: moderate** | **PA: vigorous** | **PA: summed** | **Diet** |
| Glucose | 138,200 | 138,200 | 138,200 | 138,200 | 138,200 | 138,200 | 138,200 | 116,993 |
| Haemoglobin A1C | 144,210 | 144,210 | 144,210 | 144,210 | 144,210 | 144,210 | 144,210 | 125,730 |
| Total cholesterol | 146,413 | 146,413 | 146,413 | 146,413 | 146,413 | 146,413 | 146,413 | 127,531 |
| Low density lipoprotein direct | 146,453 | 146,453 | 146,453 | 146,453 | 146,453 | 146,453 | 146,453 | 127,331 |
| High density lipoprotein cholesterol | 145,856 | 145,856 | 145,856 | 145,856 | 145,856 | 145,856 | 145,856 | 116,059 |
| C-reactive protein | 144,523 | 144,523 | 144,523 | 144,523 | 144,523 | 144,523 | 144,523 | 125,600 |
| sodium in urine | 145,241 | 145,241 | 145,241 | 145,241 | 145,241 | 145,241 | 145,241 | 129,305 |
| potassium in urine | 145,065 | 145,065 | 145,065 | 145,065 | 145,065 | 145,065 | 145,065 | 129,193 |
| Body mass index | 145,341 | 145,341 | 145,341 | 145,341 | 145,341 | 145,341 | 145,341 | 132,529 |
| Waist-hip ratio | 146,449 | 146,449 | 146,449 | 146,449 | 146,449 | 146,449 | 146,449 | 133,588 |
| Systolic blood pressure | 141,329 | 141,329 | 141,329 | 141,329 | 141,329 | 141,329 | 141,329 | 121,636 |
| Diastolic blood pressure | 141,655 | 141,655 | 141,655 | 141,655 | 141,655 | 141,655 | 141,655 | 121,931 |

To prevent the spurious signals in MRNM analysis, each of the main traits (i.e. diabetes-related metabolic traits) was pre-adjusted for confounders and excluded the outliers that three standard deviations of the mean value before applying to MRNM. The remaining participants that were used in the analysis are listed in this Table. For analyses with Age, ALC, SMK and PA, the metabolic traits were pre-adjusted for Age, ALC, SMK, PA, demographics, centre, batch and first 10 PCs. The demographic variables contain sex, year of birth, income, education, TDI. For further analysis with Diet, the metabolic traits were pre-adjusted for Age, ALC, SMK, PA, Diet, demographics, centre, batch and first 10 PCs. Therefore, the number of individuals used in the analyses with Diet were lower than that of the analyses with other covariates.

#### **Table S4. Model comparison for detecting five kinds of interaction effects**

| **Detected Interaction** |  | **Model Comparison** | **Model Equation** |
| --- | --- | --- | --- |
| 1. Overall Interaction | $H_{0}$ | MRNM null (i.e. Multivariate GREML) | $\boldsymbol{y=\mu+}\boldsymbol{\alpha}_{\mathbf{0}}\boldsymbol{+}\boldsymbol{\tau}_{\mathbf{0}}$ |
|  |  |  | $\mathbf{c}^{\boldsymbol{*}}=\boldsymbol{\mu}+\boldsymbol{\gamma}+\boldsymbol{\varepsilon}$ |
|  | $H_{1}$ | MRNM Full | $\boldsymbol{y=\mu+}\boldsymbol{\alpha}_{\mathbf{0}}\boldsymbol{+}\boldsymbol{\alpha}_{\mathbf{1}}\boldsymbol{\cdot}\mathbf{c}\boldsymbol{+}\boldsymbol{\tau}_{\mathbf{0}}\boldsymbol{+}\boldsymbol{\tau}_{\mathbf{1}}\boldsymbol{\cdot}\mathbf{c}$  $\mathbf{c}^{\boldsymbol{*}}=\boldsymbol{\mu}+\boldsymbol{\gamma}+\boldsymbol{\varepsilon}$ |
| 2. GxE interaction | $H_{0}$ | MRNM null (= Multivariate GREML) | $\boldsymbol{y=\mu+}\boldsymbol{\alpha}_{\mathbf{0}}\boldsymbol{+}\boldsymbol{\tau}_{\mathbf{0}}$  $\mathbf{c}^{\boldsymbol{*}}=\boldsymbol{\mu}+\boldsymbol{\gamma}+\boldsymbol{\varepsilon}$ |
|  | $H_{1}$ | MRNM GxE only | $\boldsymbol{y=\mu+}\boldsymbol{\alpha}_{\mathbf{0}}\boldsymbol{+}\boldsymbol{\alpha}_{\mathbf{1}}\boldsymbol{\cdot}\mathbf{c}\boldsymbol{+}\boldsymbol{\tau}_{\mathbf{0}}$  $\mathbf{c}^{\boldsymbol{*}}=\boldsymbol{\mu}+\boldsymbol{\gamma}+\boldsymbol{\varepsilon}$ |
| 3. RxE interaction | $H_{0}$ | MRNM null (= Multivariate GREML) | $\boldsymbol{y=\mu+}\boldsymbol{\alpha}_{\mathbf{0}}\boldsymbol{+}\boldsymbol{\tau}_{\mathbf{0}}$  $\mathbf{c}^{\boldsymbol{*}}=\boldsymbol{\mu}+\boldsymbol{\gamma}+\boldsymbol{\varepsilon}$ |
|  | $H_{1}$ | MRNM RxE only | $\boldsymbol{y=\mu+}\boldsymbol{\alpha}_{\mathbf{0}}\boldsymbol{+}\boldsymbol{\tau}_{\mathbf{0}}\boldsymbol{+}\boldsymbol{\tau}_{\mathbf{1}}\boldsymbol{\cdot}\mathbf{c}$  $\mathbf{c}^{\boldsymbol{*}}=\boldsymbol{\mu}+\boldsymbol{\gamma}+\boldsymbol{\varepsilon}$ |
| 4. Orthogonal RxE interaction | $H_{0}$ | MRNM GxE only | $\boldsymbol{y=\mu+}\boldsymbol{\alpha}_{\mathbf{0}}\boldsymbol{+}\boldsymbol{\alpha}_{\mathbf{1}}\boldsymbol{\cdot}\mathbf{c}\boldsymbol{+}\boldsymbol{\tau}_{\mathbf{0}}$  $\mathbf{c}^{\boldsymbol{*}}=\boldsymbol{\mu}+\boldsymbol{\gamma}+\boldsymbol{\varepsilon}$ |
|  | $H_{1}$ | MRNM Full model | $\boldsymbol{y=\mu+}\boldsymbol{\alpha}_{\mathbf{0}}\boldsymbol{+}\boldsymbol{\alpha}_{\mathbf{1}}\boldsymbol{\cdot}\mathbf{c}\boldsymbol{+}\boldsymbol{\tau}_{\mathbf{0}}\boldsymbol{+}\boldsymbol{\tau}_{\mathbf{1}}\boldsymbol{\cdot}\mathbf{c}$  $\mathbf{c}^{\boldsymbol{*}}=\boldsymbol{\mu}+\boldsymbol{\gamma}+\boldsymbol{\varepsilon}$ |
| 5. Orthogonal GxE interaction | $H_{0}$ | MRNM RxE only | $\boldsymbol{y=\mu+}\boldsymbol{\alpha}_{\mathbf{0}}\boldsymbol{+}\boldsymbol{\tau}_{\mathbf{0}}\boldsymbol{+}\boldsymbol{\tau}_{\mathbf{1}}\boldsymbol{\cdot}\mathbf{c}$  $\mathbf{c}^{\boldsymbol{*}}=\boldsymbol{\mu}+\boldsymbol{\gamma}+\boldsymbol{\varepsilon}$ |
|  | $H_{1}$ | MRNM Full model | $\boldsymbol{y=\mu+}\boldsymbol{\alpha}_{\mathbf{0}}\boldsymbol{+}\boldsymbol{\alpha}_{\mathbf{1}}\boldsymbol{\cdot}\mathbf{c}\boldsymbol{+}\boldsymbol{\tau}_{\mathbf{0}}\boldsymbol{+}\boldsymbol{\tau}_{\mathbf{1}}\boldsymbol{\cdot}\mathbf{c}$  $\mathbf{c}^{\boldsymbol{*}}=\boldsymbol{\mu}+\boldsymbol{\gamma}+\boldsymbol{\varepsilon}$ |

Five kinds of model comparisons used in this study were represented based on the description of MRNM (see Note S3). The likelihood ratio values were obtained to test whether the alternative hypothesis ($H_{1}$) is much likely to occur compared to the null ($H_{0}$).

#### **Table S5. Linear regression coefficients between T2DM and 12 diabetes-related metabolic traits**

| **Related Traits** | **Abbreviations** | **# in Traits** | **R^2^ (%)** | **p-value** |
| --- | --- | --- | --- | --- |
| HemoglobinA1c | HbA1c | 274,574 | 23.500 | 2.20E-16 |
| Blood Glucose | Glucose | 251,220 | 15.000 | 2.20E-16 |
| Total Cholesterol | TC | 274,640 | 4.757 | 2.20E-16 |
| Low density lipoprotein cholesterol | LDL | 274,135 | 4.193 | 2.20E-16 |
| Body mass index | BMI | 287,322 | 3.767 | 2.00E-16 |
| Waist-Hip ratio | WHR | 287,694 | 3.208 | 2.00E-16 |
| High density lipoprotein cholesterol | HDL-C | 251,398 | 2.131 | 2.00E-16 |
| C-reactive Protein | CRP | 274,054 | 0.161 | 2.20E-16 |
| Sodium in Urine | Sodium | 279,344 | 0.160 | 2.20E-16 |
| Systolic Blood Pressure | Systolic BP | 262,734 | 0.145 | 2.20E-16 |
| Potassium in Urine | Potassium | 279,329 | 0.046 | 2.20E-16 |
| Diastolic Blood Pressure | Diastolic BP | 262,739 | 0.025 | 3.24E-16 |

The associations between 12 diabetes-related metabolic traits and T2DM disease status were tested using a linear model to explore how much they are related

#### **Table S6. Estimates with significant signals for orthogonal GxE interaction from the meta-analysed results.**

| **Main Trait** | **Lifestyle covariate** | $\boldsymbol{\sigma}_{\boldsymbol{\tau}_{\boldsymbol{0}}}^{\boldsymbol{2}}$ | $\boldsymbol{\sigma}_{\boldsymbol{\tau}_{\boldsymbol{1}}}^{\boldsymbol{2}}$ | $\boldsymbol{\sigma}_{\boldsymbol{\tau}_{\boldsymbol{0}}\boldsymbol{,}\boldsymbol{\tau}_{\boldsymbol{1}}}$ | $\boldsymbol{\sigma}_{\boldsymbol{g}_{\boldsymbol{0}}}^{\boldsymbol{2}}$ | $\boldsymbol{\sigma}_{\boldsymbol{g}_{\boldsymbol{1}}}^{\boldsymbol{2}}$ | $\boldsymbol{\sigma}_{\boldsymbol{g}_{\boldsymbol{0}}\boldsymbol{,}\boldsymbol{g}_{\boldsymbol{1}}}$ |
| --- | --- | --- | --- | --- | --- | --- | --- |
| Glucose | Alcohol consumption | 0.905(0.003) | 0.019 (0.003) | 0.021 (0.002) | 0.074 (0.002) | 0.003 (0.002) | 0.000 (0.002) |
|  | Physical activity: walk | 0.901 (0.003) | 0.030 (0.003) | -0.032 (0.002) | 0.075 (0.002) | 0.007 (0.002) | -0.004 (0.002) |
| HbA1c | Physical activity: moderate | 0.776 (0.003) | 0.008 (0.002) | -0.022 (0.002) | 0.207 (0.003) | 0.009 (0.002) | -0.005 (0.002) |
| Total Cholesterol | Age at recruitment | 0.848 (0.003) | 0.026 (0.003) | 0.060 (0.002) | 0.114 (0.002) | 0.016 (0.002) | -0.008 (0.002) |
| BMI | Age at recruitment | 0.808 (0.003) | -0.026 (0.003) | -0.015 (0.002) | 0.206 (0.003) | 0.009 (0.002) | -0.013 (0.002) |
|  | Alcohol consumption | 0.740 (0.003) | 0.040 (0.003) | 0.059 (0.002) | 0.208 (0.003) | 0.009 (0.002) | 0.029 (0.002) |
|  | Smoking Status | 0.776 (0.003) | -0.008 (0.003) | 0.020 (0.002) | 0.207 (0.003) | 0.025 (0.003) | -0.002 (0.002) |
|  | Diet | 0.789 (0.003) | -0.007 (0.003) | -0.008 (0.002) | 0.213 (0.003) | 0.005 (0.002) | -0.011 (0.002) |
| Waist-Hip ratio | Alcohol consumption | 0.829 (0.003) | 0.009 (0.003) | 0.024 (0.002) | 0.152 (0.002) | 0.010 (0.002) | 0.016 (0.002) |
| Systolic BP | Smoking Status | 0.842 (0.003) | -0.01 (0.003) | 0.020 (0.002) | 0.155 (0.003) | 0.013 (0.003) | -0.012 (0.002) |
|  | Physical activity: moderate | 0.854 (0.003) | -0.015 (0.003) | 0.019 (0.002) | 0.155 (0.003) | 0.006 (0.002) | 0.000 (0.002) |
| Diastolic BP | Smoking Status | 0.850 (0.003) | -0.005 (0.002) | 0.011 (0.002) | 0.146 (0.003) | 0.008 (0.002) | -0.014 (0.002) |
| LDL Cholesterol | Age at recruitment | 0.868 (0.003) | 0.022 (0.003) | 0.057 (0.002) | 0.099 (0.002) | 0.015 (0.002) | -0.008 (0.002) |
| HDL cholesterol | Diet | 0.740 (0.004) | -0.001 (0.003) | 0.023 (0.002) | 0.258 (0.003) | 0.008 (0.003) | 0.012 (0.002) |
| C reactive protein | Alcohol consumption | 0.798 (0.003) | 0.055 (0.003) | 0.074 (0.002) | 0.135 (0.002) | 0.006 (0.002) | 0.010 (0.002) |
|  | Physical activity: summed | 0.843 (0.003) | 0.011 (0.002) | -0.064 (0.002) | 0.137 (0.002) | 0.007 (0.002) | -0.016 (0.001) |
|  | Physical activity: moderate | 0.845 (0.003) | 0.015 (0.002) | -0.052 (0.002) | 0.137 (0.002) | 0.004 (0.002) | -0.011 (0.002) |

### Supplementary Figures

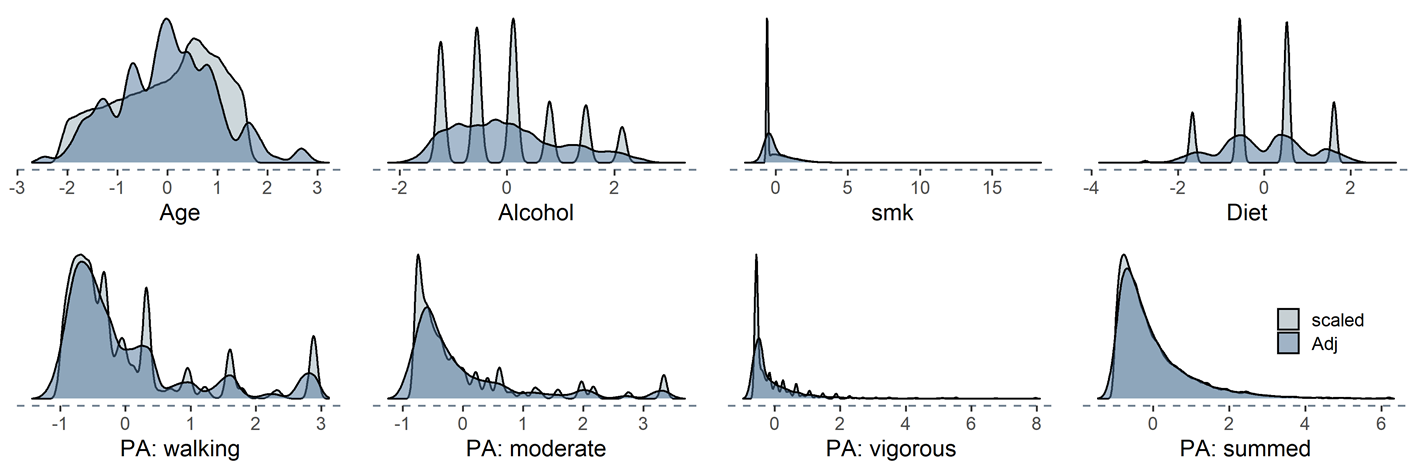

#### **Figure S1. Histograms for the lifestyle covariate that showed their distribution of scaled only and pre-adjusted.**

The distribution of 8 lifestyles that were applied as the second trait and covariate in the model were plotted. We used the pre-adjusted covariates when the lifestyle is fitted as the second trait (Adj), and standardised covariates were applied as covariates in the model (scaled). Noting that the pre-adjusted covariates are also the standardised values after the adjustment.

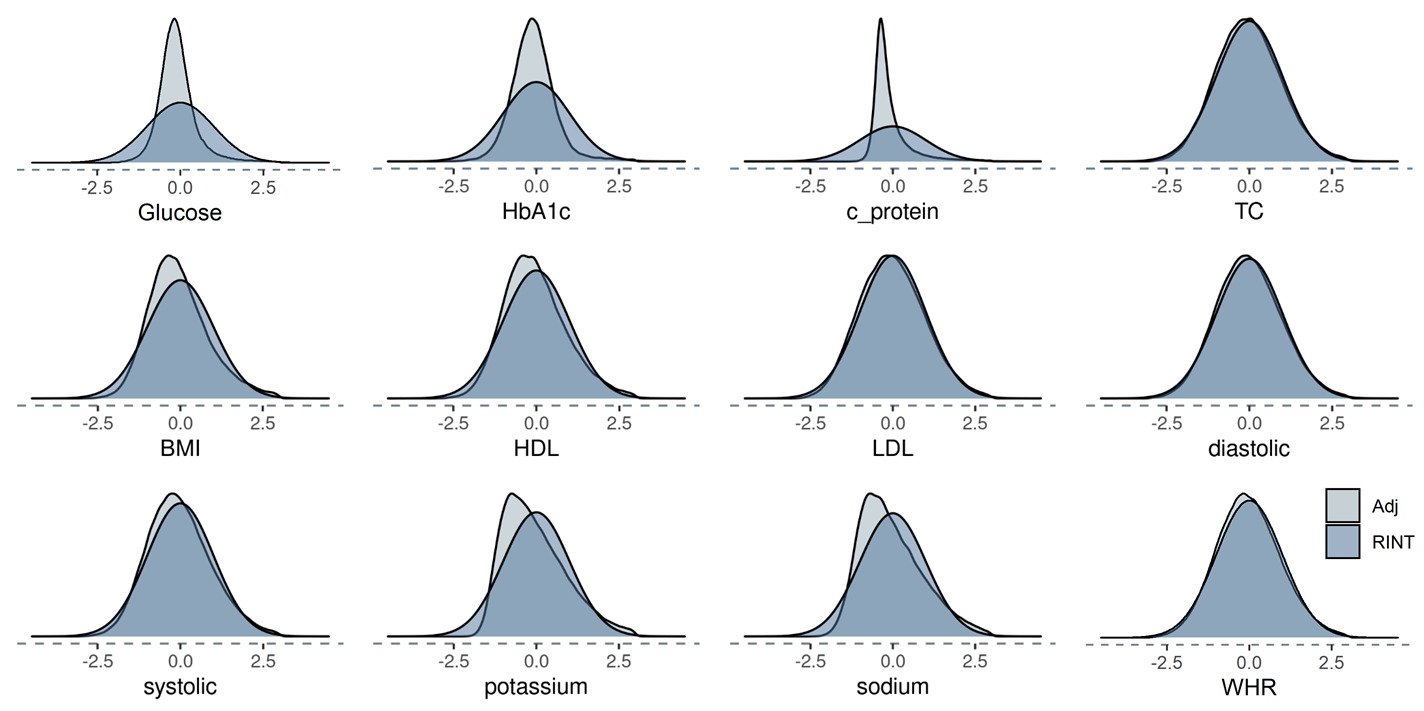

#### **Figure S2. Histograms for the diabetes-related metabolic traits that showed their distribution before and after applying RINT.**

Diabetes-related metabolic traits, which are used as the main traits in MRNM analysis, were applied a rank-based inverse normal transformation (RINT) to avoid the possibility of obtaining false signals from the usage of data with large skewness and kurtosis. The figures showed the distribution change before and after application of RINT, where Adj (light blue) indicates the trait that pre-adjusted for the covariates and excluded both 3SD outer sides, and RINT (dark blue) indicates that RINT is applied to the Adj traits.

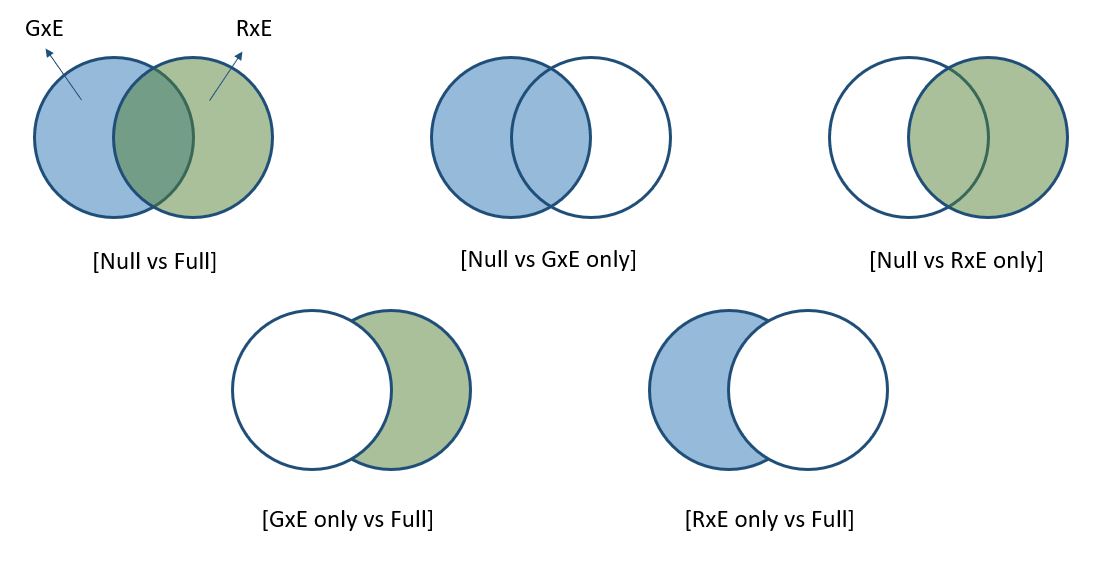

#### **Figure S3. The illustration of five different model comparisons.**

Likelihood ratio tests were used to find a better model and the model comparisons involve 4 different models, which are null, GxE only, RxE only and full models. The blue or green colour indicates the part of GxE or RxE interaction. The design that is compared null and full models is for identifying the overall GxE and RxE interactions, compared GxE only and full models is for the orthogonal RxE interaction and compared RxE only and full models is for the orthogonal GxE interaction.

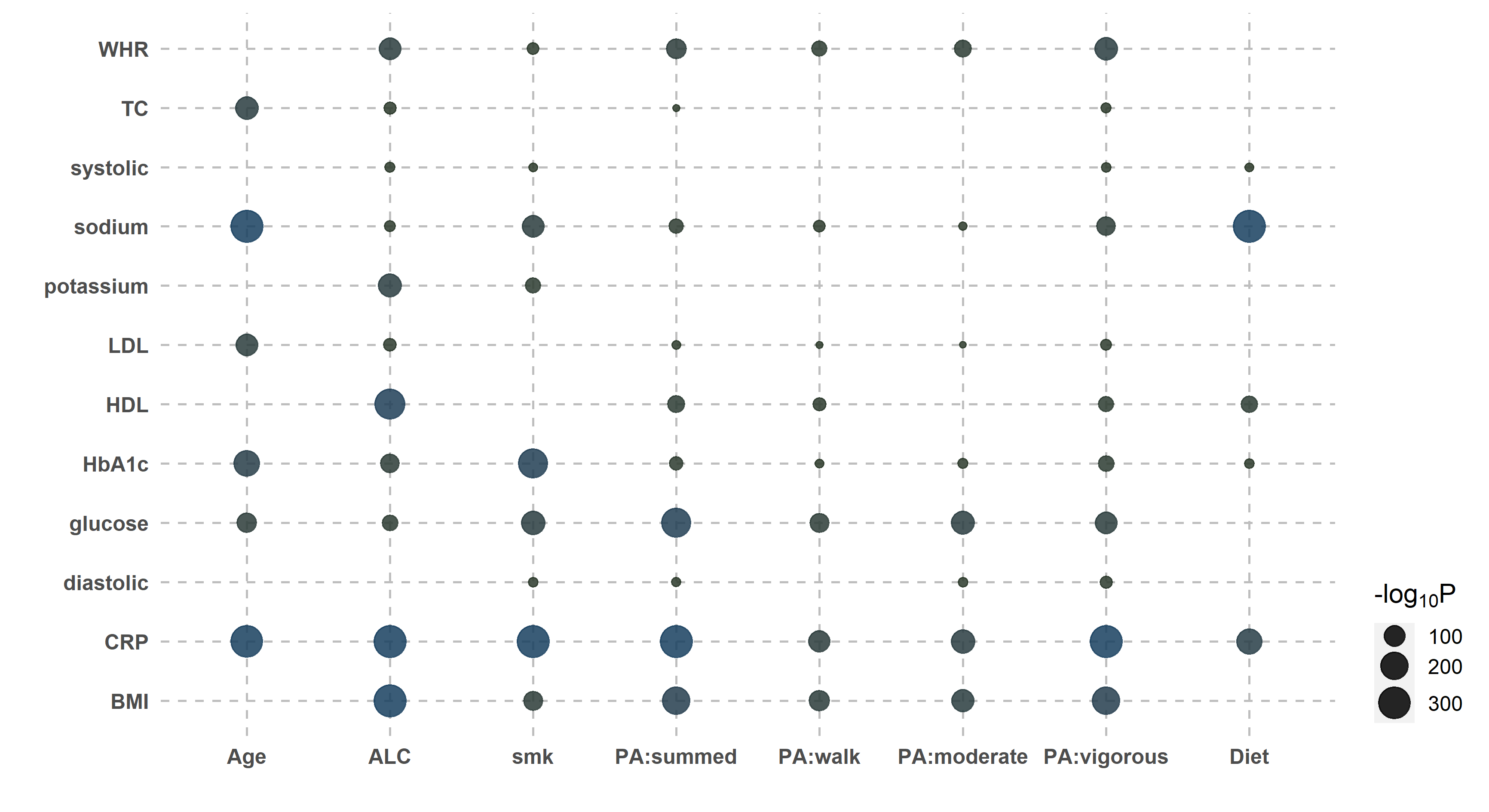

#### **Figure S4. Bubble plot of p-values indicating significant overall interactions (i.e. GxE and/or RxE).**

There were 68 significant overall interactions when testing the genetic and non-genetic effects of 12 diabetes-related metabolic traits modulated by 8 covariates. Likelihood ratio tests were used to compare the full model with null model for each of 6 independent datasets, and p-values were meta-analysed using the Fisher’s method. The size of dot reflects its significance, the bigger the more significant.

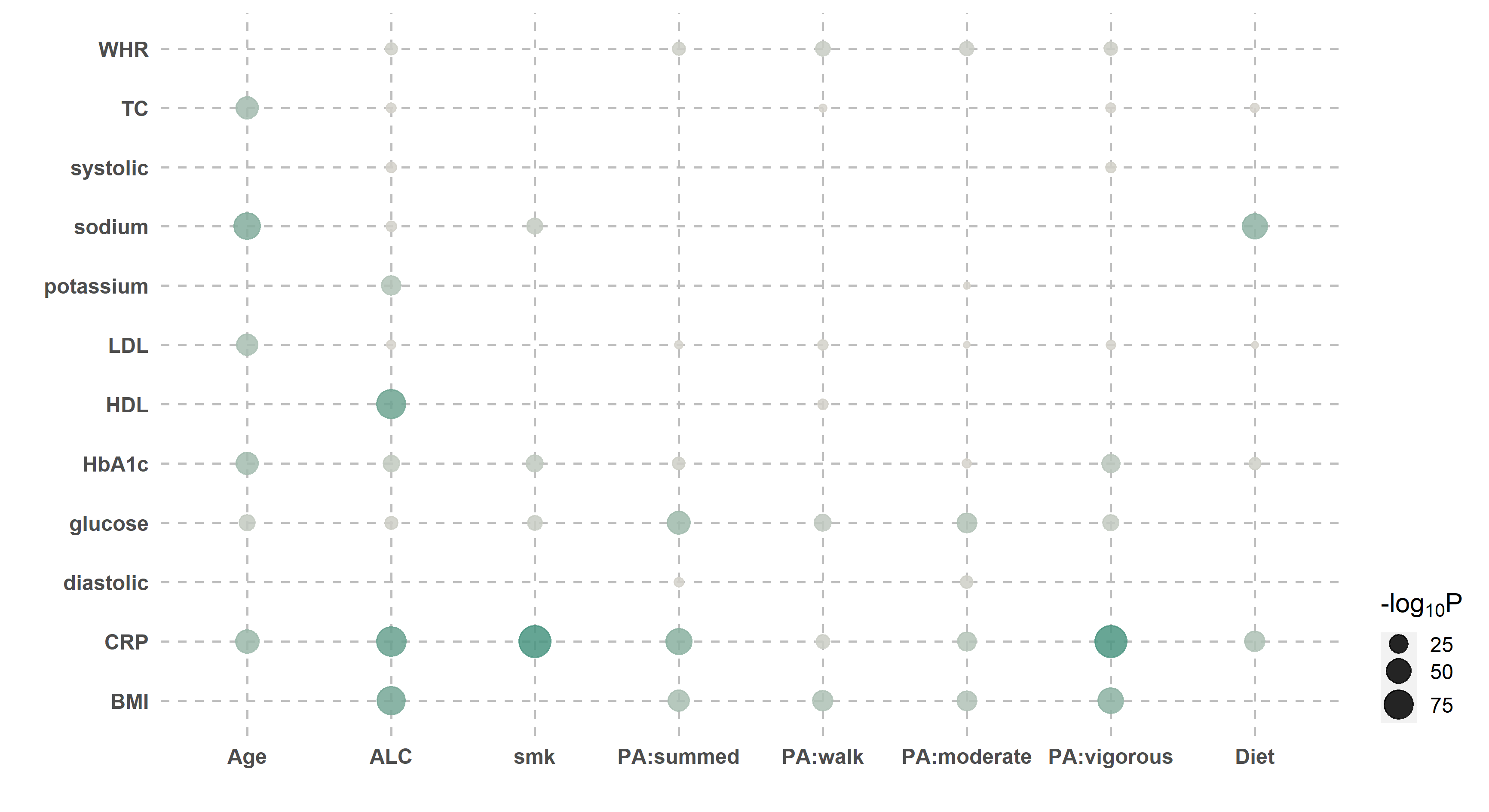

#### **Figure S5. Bubble plot of p-values indicating significant orthogonal RxE interactions.**

There were 56 significant orthogonal RxE interactions when testing the non-genetic effects of 12 diabetes-related metabolic traits modulated by 8 covariates. Likelihood ratio tests were used to compare the full model with GxE only model for each of 6 independent datasets, and p-values were meta-analysed using the Fisher’s method. The size of dot reflects its significance, the bigger the more significant.

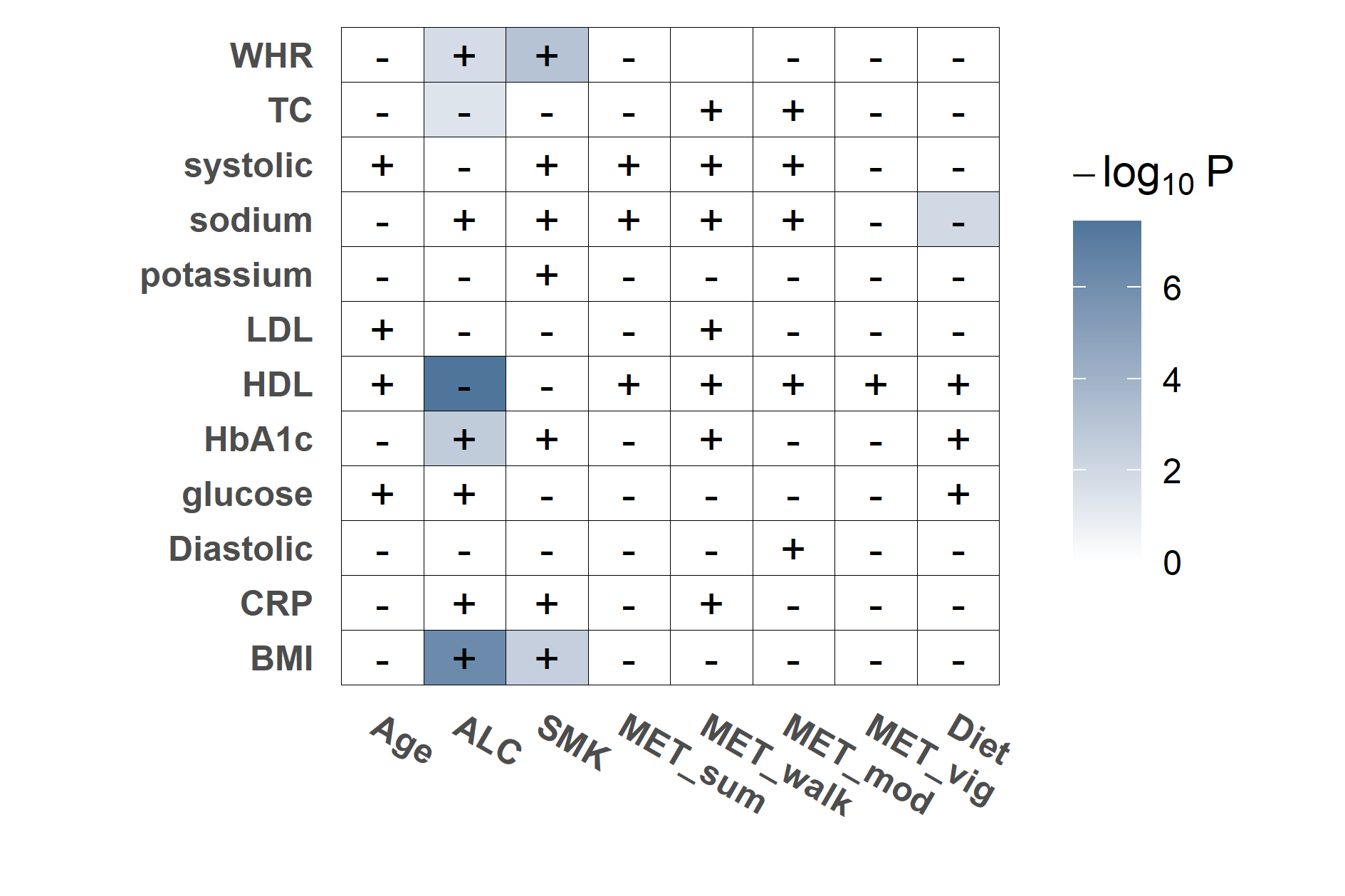

#### **Figure S6. Heatmap plot for the causality.**

Coloured tiles indicate the significant signal for a causal effect of the environmental covariates (columns) on the main traits (rows), given the estimated p-value testing that casual model is a better fit (p-values > 0.05). The white tiles indicate that there is no significant causal effect (p-values <0.05). The signs on each tile indicate (i.e., plus and minus) the direction of effects. A plus sign means that the level of main traits increases when the level of environmental covariates increases, and conversely, the main traits with the negative sign decrease.

*MET_sum; total physical activity, MET_vig; vigorous physical activity, MET_mod; moderate physical activity, MET_walk; walking physical activity, SMK; smoking status, ALC; alcohol intake frequency, BMI; body mass index, CRP; C-Reactive protein, HbA1c; Hemoglobin A1C, WHR; waist-hip ratio, TC; total cholesterol, Diastolic; Diastolic blood pressure, Systolic; Systolic blood pressure, HDL; high density lipoprotein cholesterol, LDL; low density lipoprotein cholesterol, sodium; sodium in urine, potassium; potassium in urine.*

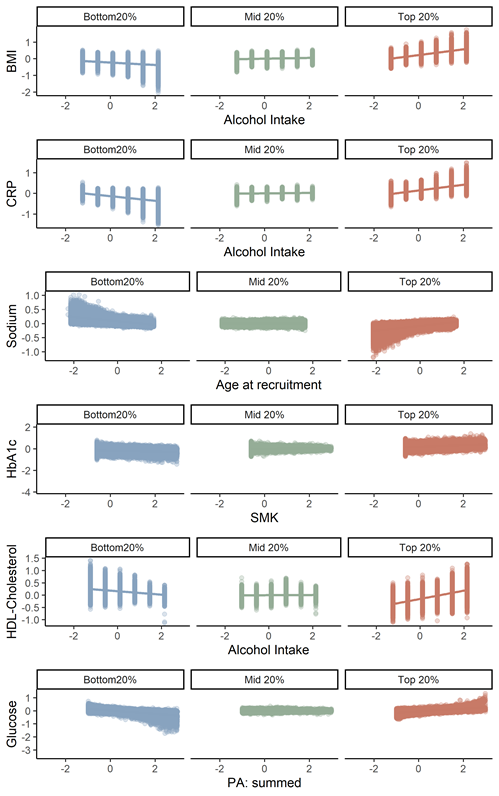

#### **Figure S7. Individuals phenotypic effect of six traits with the most significant overall interaction in response to the level of environmental covariates.**

The phenotypic effect for each individual was estimated by reflecting both GxE and RxE interaction (i.e.,$\hat{\mathbf{y}}$, see methods), and the phenotypic effects of the Bottom 20% (blue), Middle 20% (green) and Top20% (red) groups for the estimated orthogonal interaction effects are represented, showing the aspect that changes depending on the level of covariates. The limit of the scale for the covariate (x axis) is set from -3 to 3.

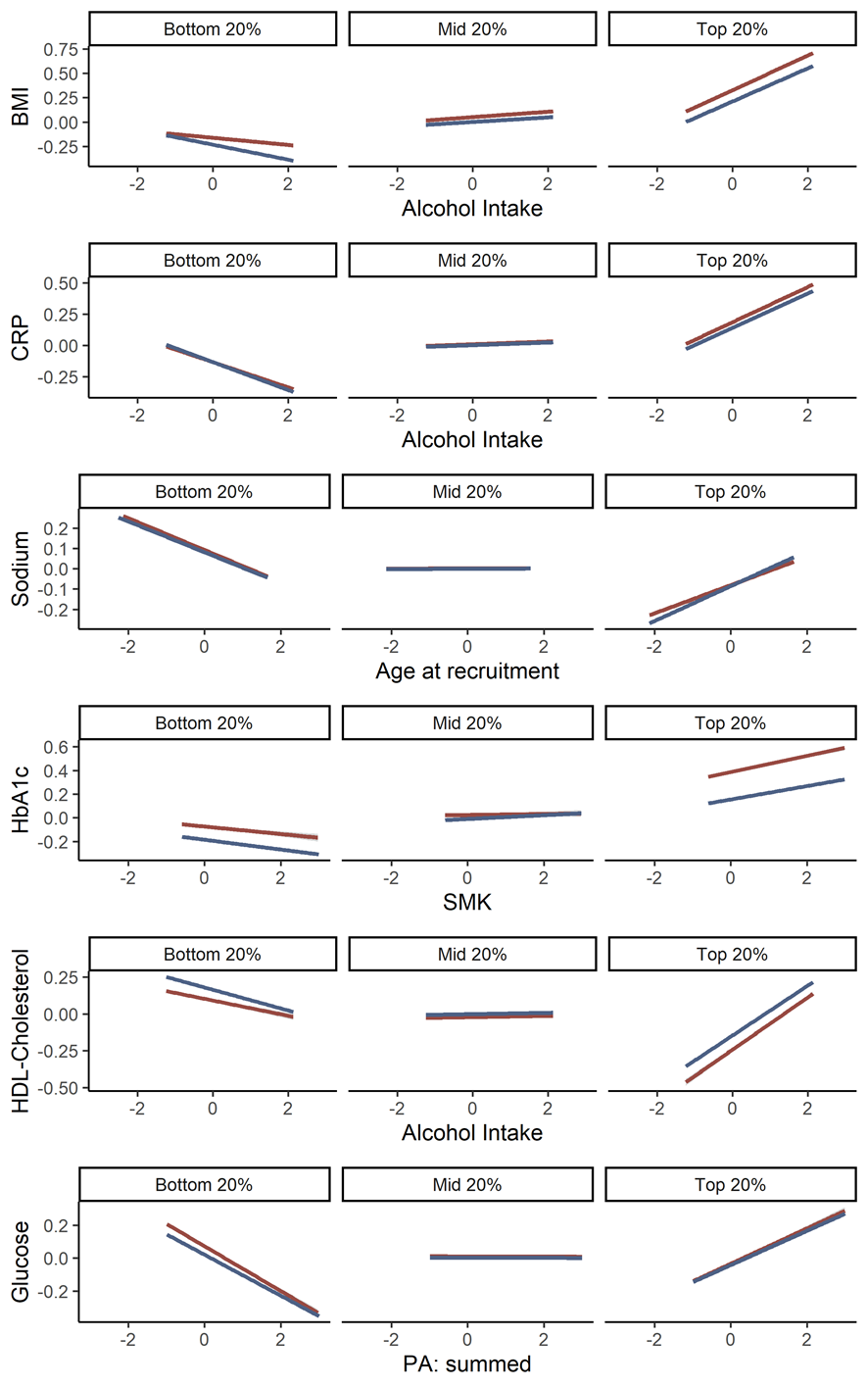

#### **Figure S8. Plasticity of six traits with the most significant overall interaction in response to the level of environmental effects, by grouping T2DM prospective cases and controls.**

To compare the differences in trajectories between the participants with T2DM prospective cases (red) and controls (blue), linear regressions between the main trait and covariate were represented for 6 traits with most significant overall interaction, and we further grouped into the bottom 20%, middle 20% and top 20% depending on the estimated combined interaction (i.e. $\sigma_{\alpha_{1}}^{2}$+$\sigma_{\tau_{1}}^{2}$). The phenotype for each trait can be written as $\hat{\mathbf{y}}=\hat{\boldsymbol{\alpha}_{\mathbf{0}}}+\hat{\boldsymbol{\alpha}_{\mathbf{1}}}\cdot\mathbf{c+}\hat{\boldsymbol{\tau}_{\mathbf{1}}}\cdot\mathbf{c}$ (see Methods). Noting that the direction is not considered, i.e. it goes to the favourable when the level of physical activity increases. The limit of the scale for the covariate (x axis) is set from -3 to 3.

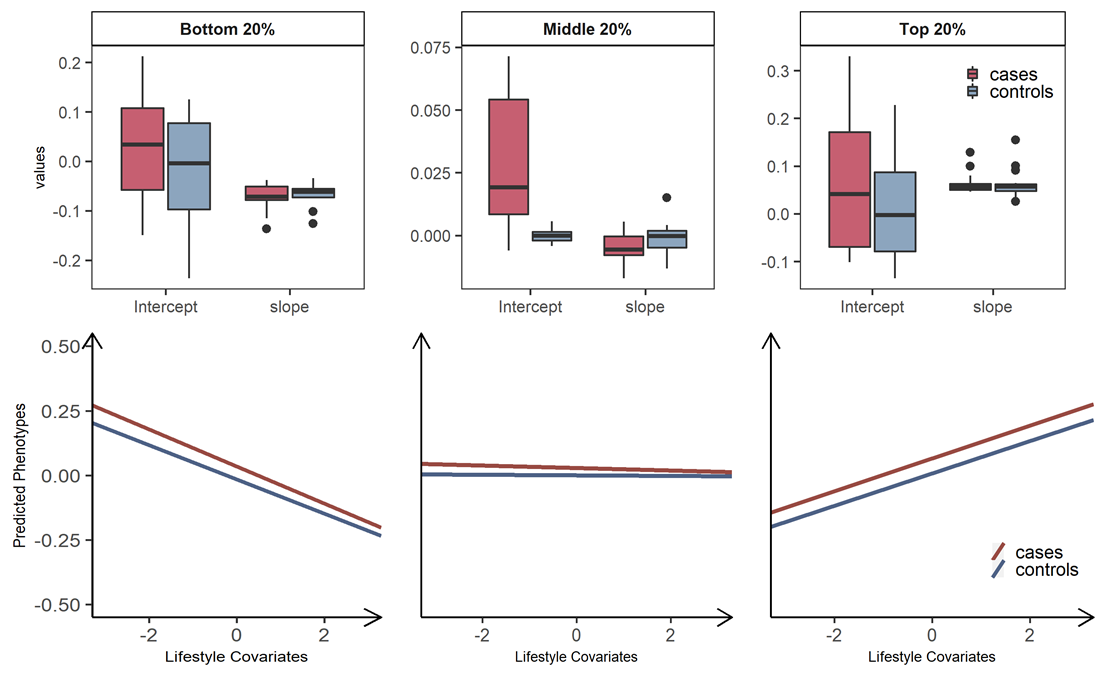

#### **Figure S9. Plasticity of diabetes-related metabolic traits of 17 significant GxE interactions in response to the level of lifestyle effects, by grouping T2DM prospective cases and controls.**

Individuals are stratified into three groups, the bottom, middle and top 20% groups according to estimated GxE interaction from the full model (i.e. $\sigma_{\alpha_{1}}^{2}$). We further classified individuals into T2DM prospective cases (red) and controls (blue) in each of three groups, hence six groups in total. To compare the differences in trajectories between with T2DM cases and controls, a linear model regressing the phenotype of main trait on standardised lifestyle measures was used to estimate the intercept and slope for each of six groups. The values of intercepts and slopes were averaged over the 17 analyses that showed significant signals for orthogonal GxE interactions. The averaged values of intercepts and slopes represent the overall relationship between metabolic risk and lifestyle covariates, where we considered making favourable and unfavourable directions consistent across the main traits and covariates, to facilitate a better interpretation in line with metabolic risks on T2DM (see Methods). Box plot for each group was represented to show the differences in terms of the calculated intercepts and slopes, and the mean of values for intercepts and slopes were represented as regression lines. The mean and standard error of intercepts and slopes across the analyses of the 17 pairs with significant overall interactions were estimated to assess if there is any significant difference of the mean between cases and control in each of the three groups, the bottom, middle and top 20% groups. There were no significant differences between cases and controls in the mean of slopes in all three groups. The mean of intercepts is significantly different between cases and controls in all three groups (p-values= 1.9E-03, 9.47E-05 and 3.54E-04). The arrows on both axes in linear regression figures indicate an unfavourable direction in regard to T2DM health.

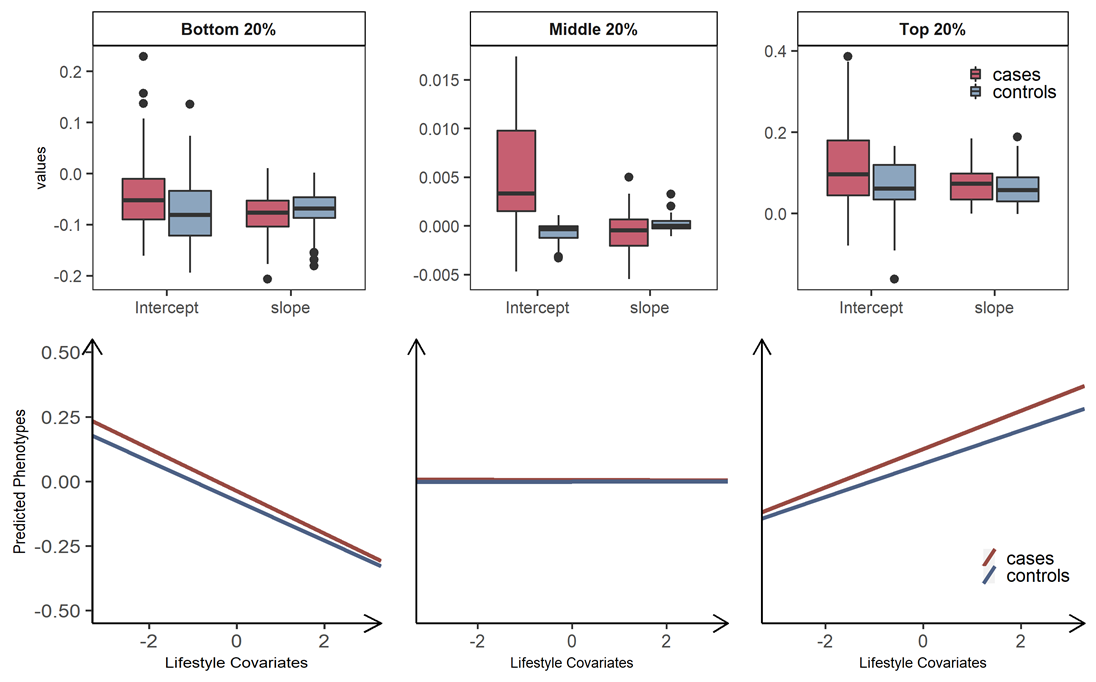

#### **Figure S10. Plasticity of diabetes-related metabolic traits of 56 significant RxE interactions in response to the level of lifestyle effects, by grouping T2DM prospective cases and controls.**

Individuals are stratified into three groups, the bottom, middle and top 20% groups according to estimated RxE interaction from the full model (i.e.$\sigma_{\tau_{1}}^{2}$). We further classified individuals into T2DM prospective cases (red) and controls (blue) in each of three groups, hence six groups in total. To compare the differences in trajectories between with T2DM cases and controls, a linear model regressing the phenotype of main trait on standardised lifestyle measures was used to estimate the intercept and slope for each of six groups. The values of intercepts and slopes were averaged over the 56 analyses that showed significant signals for orthogonal RxE interactions. The averaged values of intercepts and slopes represent the overall relationship between metabolic risk and lifestyle covariates, where we considered making favourable and unfavourable directions consistent across the main traits and covariates, to facilitate a better interpretation in line with metabolic risks on T2DM (see Methods). Box plot for each group was represented to show the differences in terms of the calculated intercepts and slopes, and the mean of values for intercepts and slopes were represented as regression lines. The mean and standard error of intercepts and slopes across the analyses of the 56 pairs with significant overall interactions were estimated to assess if there is any significant difference of the mean between cases and control in each of the three groups, the bottom, middle and top 20% groups. The mean of slopes in incidence cases significantly differs from controls in all three groups (p-values=6.60E-03, 5.68E-03 and 1.41E-05). The mean of intercepts is significantly different between cases and controls in all three groups (p-values= 6.03E-05, 1.97E-10 and 7.71E-07). The arrows on both axes in linear regression figures indicate an unfavourable direction in regard to T2DM health.

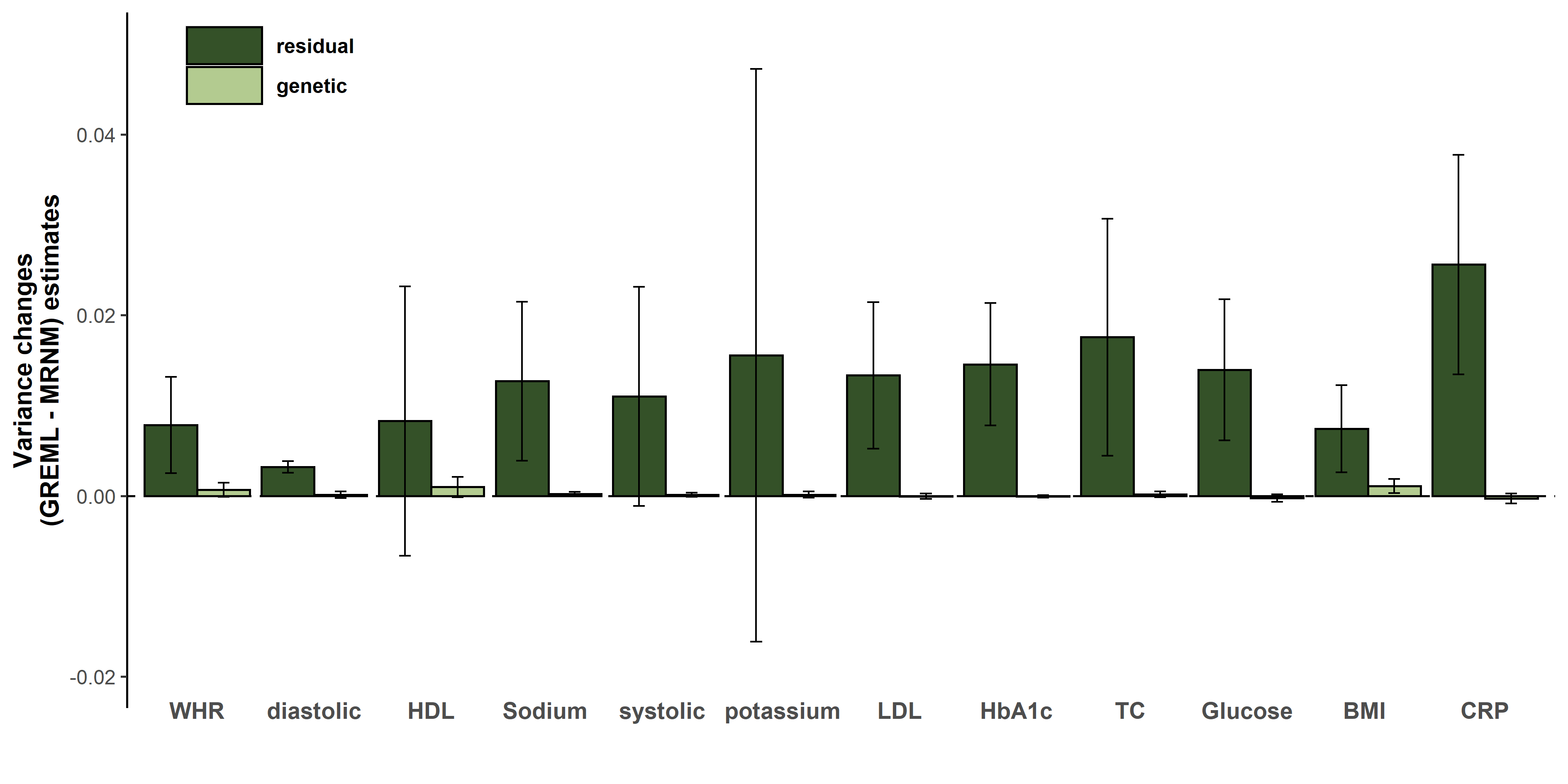

#### **Figure S11. Variance differences between the bivariate GREML and MRNM analyses in genetic and non-genetic effects.**

The bars indicate the main genetic and residual variance differences, i.e., ($\sigma_{\alpha_{0}\_GREML}^{2}-\sigma_{\alpha_{0}\_MRNM}^{2}$) and ($\sigma_{\tau_{0}\_GREML}^{2}-\sigma_{\tau_{0}\_MRNM}^{2}$). Where $\sigma_{\alpha_{0}\_GREML}^{2}$ and $\sigma_{\alpha_{0}\_MRNM}^{2}$ are the estimated main genetic variances from the GREML and MRNM, and $\sigma_{\tau_{0}\_GREML}^{2}$ and $\sigma_{\tau_{0}\_MRNM}^{2}$ are the main residual variances obtained from the GREML and MRNM. The vertical line of each bar is the 95% confidence interval for the variance changes. Thus, the positive bars indicate that the values of GREML are larger than that of MRNM, and the negative bars mean that the values of MRNM are larger than that of MRNM. The greater changes in residuals than that of the genetics may imply that most proportion of interactions were disentangled from the main residuals, not genetics. 68 pairs of significant overall interacions were used in the comparisons and noting that a single environmental covariat is considered for each pair (i.e. not fitting multiple covariates).

*WHR; waist-hip ratio, diastolic; diastolic blood pressure, HDL; high density lipoprotein cholesterol, sodium; sodium in urine, LDL; low density lipoprotein cholesterol, potassium; potassium in urine, TC; total cholesterol, HbA1c; hemoglobinA1C, BMI; body mass index, CRP; C reactive protein, systolic; systolic blood pressure.*
